# Estimating the Effect of PrEP in Black Men Who Have Sex with Men: A Framework to Utilize Data from Multiple Non-Randomized Studies to Estimate Causal Effects

**DOI:** 10.1101/2024.01.10.24301113

**Authors:** Allison Meisner, Fan Xia, Kwun C.G. Chan, Kenneth Mayer, Darrell Wheeler, Sahar Zangeneh, Deborah Donnell

**Author notes:** Address: 1100 Fairview Ave N, M4-B829, Seattle, WA, US 98109.

## Abstract

Black men who have sex with men (MSM) are disproportionately burdened by the HIV epidemic in the US. The effectiveness of pre-exposure prophylaxis (PrEP) in preventing HIV infection has been demonstrated through randomized placebo-controlled clinical trials in several populations. Importantly, no such trial has been conducted exclusively among Black MSM in the US, and it would be unethical and infeasible to do so now. To estimate the causal effects of PrEP access, initiation, and adherence on HIV risk, we utilized causal inference methods to combine data from two non-randomized studies that exclusively enrolled Black MSM. The estimated relative risks of HIV were: (i) 0.52 (95% confidence interval: 0.21, 1.22) for individuals with versus without PrEP access, (ii) 0.48 (0.12, 0.89) for individuals who initiated PrEP but were not adherent versus those who did not initiate, and (iii) 0.23 (0.02, 0.80) for individuals who were adherent to PrEP versus those who did not initiate. Beyond addressing the knowledge gap around the effect of PrEP in Black MSM in the US, which may have ramifications for public health, we have provided a framework to combine data from multiple non-randomized studies to estimate causal effects, which has broad utility.

## INTRODUCTION

The HIV epidemic, now in its fifth decade, has been concentrated in specific key populations since it began. One group that has been disproportionately affected since the very start of the epidemic is men who have sex with men (MSM). In the US, MSM accounted for 71% of all new HIV diagnoses in 2020 [1]. Among MSM in the US, Black men are at particular risk, accounting for 26% of all new HIV diagnoses and 39% of diagnoses among MSM in 2020 [1] despite Black people comprising only 13.6% of the US population [2]. It is believed that Black MSM are at high risk of HIV due to a higher prevalence of sexually transmitted infections (STIs), higher levels of unrecognized HIV in their sexual networks (which has downstream effects such as delayed initiation of anti-retroviral therapy, a treatment which can reduce the risk of transmission of HIV), sex partner demographics, and a constellation of marginalizing factors, including lower income, underemployment, educational inequalities, inadequate access to healthcare, incarceration, stigma, and discrimination [3-10].

The high risk and disproportionate burden of HIV among Black MSM persisted even after pre-exposure prophylaxis (PrEP) was approved by the Food and Drug Administration in 2012. PrEP has been shown to be highly effective in preventing HIV infection in a variety of populations, including MSM [11-18]. However, while previous trials have included Black MSM in the US, they have not been sufficiently represented in randomized, placebo-controls trials to date. Consequently, little is known about the effectiveness of PrEP specifically within the unique context of this key population. Given the extensive evidence that PrEP is effective in general, it is not ethical to randomize Black MSM to placebo. Thus, a knowledge gap regarding the effectiveness of PrEP in this vulnerable population persists. Quantifying the effectiveness of PrEP in this population could provide insights into the generalizability of effectiveness estimates from other populations, highlighting factors which may modify the effect of PrEP. From a public health perspective, an estimate of effectiveness specific to Black MSM may motivate Black MSM to initiate and adhere to PrEP and may encourage their clinicians to support such behavior, leading to reductions in HIV incidence.

We sought to estimate the effect of PrEP on HIV risk in Black MSM in the US by combining data from two large studies conducted exclusively in Black MSM by the HIV Prevention Trials Network (HPTN), including a single-arm study and an observational study. We focused on the effect of PrEP on HIV risk at three stages of the PrEP engagement “cascade”, namely, the effect of PrEP access, the effect of PrEP initiation without adherence, and the effect of PrEP adherence. To estimate causal effects from non-randomized data while leveraging data from a single-arm study, we utilized methods from causal inference, including inverse probability weighting (IPW) [19, 20]. Consequently, beyond estimating the effectiveness of PrEP in reducing risk of HIV in Black MSM in the US, our work provides a general framework to estimate causal effects using data from multiple, non-randomized studies.

## METHODS

### Data Sources

Data from two studies in HPTN were used in this analysis: HPTN 061 and HPTN 073. HPTN 061 was an observational study of Black MSM in several cities in the US which sought to evaluate the feasibility and acceptability of a multi-component HIV prevention intervention [21]. The study was conducted between 2009 and 2011; thus, we assumed that there was functionally no access to PrEP for men in this study as the study ended prior to PrEP approval. The study enrolled 1,162 men who did not have HIV, 28 of whom were known to have acquired HIV during one year of follow-up [21]. The Supplement includes additional information on the design of HPTN 061.

HPTN 073 was a non-randomized, open-label PrEP study of Black MSM in several US cities [22]. The study was conducted between 2013 and 2015, i.e., immediately after FDA approval of PrEP, but close in time to HPTN 061. Consequently, all men enrolled in HPTN 073 had access to PrEP through participation in the study (daily oral co-formulated emtricitabine and tenofovir disoproxil fumarate (FTC/TDF)) and could decide whether to initiate and/or adhere to PrEP at any point. Importantly, access to PrEP in HPTN 073 was of an “active” form: beyond PrEP being available (i.e., “passive access”), participants in HPTN 073 had frequent study visits wherein PrEP was offered and study staff were available to assist participants if they expressed an interest in initiating. A total of 226 men without HIV were enrolled and followed for one year. For those men who initiated PrEP (nearly 80%), the date of first dose was recorded. Blood samples were subsequently taken to measure PrEP adherence, defined as at least 4 doses per week [23]. Eight participants were known to have acquired HIV during one year of follow-up [22]. The Supplement includes additional information on the design of HPTN 073.

In order to use data from both HPTN 061 and HPTN 073, the cohorts, timing of measurements, and variables assessed needed to be as similar as possible; see the Supplement for full details. Measurements (including those of relevant covariates and HIV status) were more frequent in HPTN 073 than in HPTN 061 (quarterly vs. biannually), so we utilized the data from the enrollment, 6-month, and 12-month visits in both studies. We also restricted attention to those variables measured in both studies.

Using the nomenclature of Degtiar and Rose [24], we define our *target population*, i.e., that in which we would like to make inference, as Black MSM in the US. Our *study population*, in which the effect will actually be estimated and inference made, is the HPTN 073 population, that is, the population from which the HPTN 073 study participants were (randomly) sampled. We chose the HPTN 073 population to be our study population since it is more recent. Our analysis was based on two study samples, the HPTN 073 sample, which is representative of the HPTN 073 population, and the HPTN 061 sample, which is representative of the HPTN 061 population.

### Analysis

Our goal was to estimate the effects of PrEP access, initiation without adherence (“initiation” hereafter), and adherence on HIV risk in Black MSM in the US. We sought to estimate the marginal effect of each of these exposures, i.e., not conditional on any covariates, consistent with the effect that would be estimated in a randomized trial. Let *A* denote PrEP access: and *A*= 1 indicates no PrEP access and *A* = 0 indicates no PrEP access. The variable *D* denotes PrEP initiation and adherence: *D* = 0 if the participant has not initiated, (and thus is not adherent), *D* = 1 if the participant has initiated but is not adherent, and *D* = 2 if the participant has initiated and is adherent. In HPTN 061, *A* = *D* = 0. In HPTN 073, *A* = 1 and *D* was 0, 1, or 2. We define *Y* to be HIV status (*Y*= 1 if the participant has HIV and *Y*= 0 if they do not) and ***X*** is a vector of baseline and time-varying covariates.

Both studies collected data at 0, 6, and 12 months; these time points are denoted by *t* = 0,1,2, respectively. We conceptualize the time between 0 and 6 months as the “first interval” and the time between 6 and 12 months as the “second interval”. Access (*A*) is time-invariant. The covariates ***X***_***t***_ denote the vector of covariate values measured at time t (including both time varying and time-invariant covariates). Likewise, *D*_*t*_ denotes PrEP initiation/adherence assessed at time *t, t* =1,2 ; e.g., *D*_*t*_ = 1 if the participant initiated PrEP prior to time *t* but was not adherent at time *t*. Finally, *Y*_1_ and *Y*_2_ indicate whether the participant had HIV by *t* = 1 and *t* = 2, respectively. We use the subscript *i* to refer to values of the exposures, covariates, and outcomes for individual *i, i* = 1 …, *n*, where *n = n*_061_ + *n*_073_, the total number of participants in the HPTN 061 and HPTN 073 samples.

A directed acyclic graph (DAG) characterizing the relationships between the variables is given in Figure 1. Dashed lines are drawn from *A* to *D*_1_ and *D*_2_ because there is a (partially) deterministic relationship at play: if *A* = 0, then *D*_1_ = *D*_2_ = 0. Likewise, there are dashed lines drawn from *Y*_1_ to *D*_2_ and *Y*_2_ because there is a (partially) deterministic relationship between these variables as well: if *Y*_1_ = 1, then *Y*_2_ = 1 and *D*_2_ is not relevant (a person would not keep taking PrEP if they had previously tested positive for HIV). We suppose that PrEP access (*A*) has no direct effect on HIV risk (*Y*_1_ and *Y*_2_) and the effect of access is only through PrEP initiation and adherence. Some have argued that taking PrEP influences the probability of engaging in higher risk behaviors (which may lead to an arrow from *D*_1_ to ***X***_1_, since ***X***_1_ likely includes some of these behaviors); however, the preponderance of evidence indicates that any effect is of minimal magnitude [25, 26]. Thus, we assume there is no causal link between *D*_1_ and ***X***_1_.

**Figure 1:**
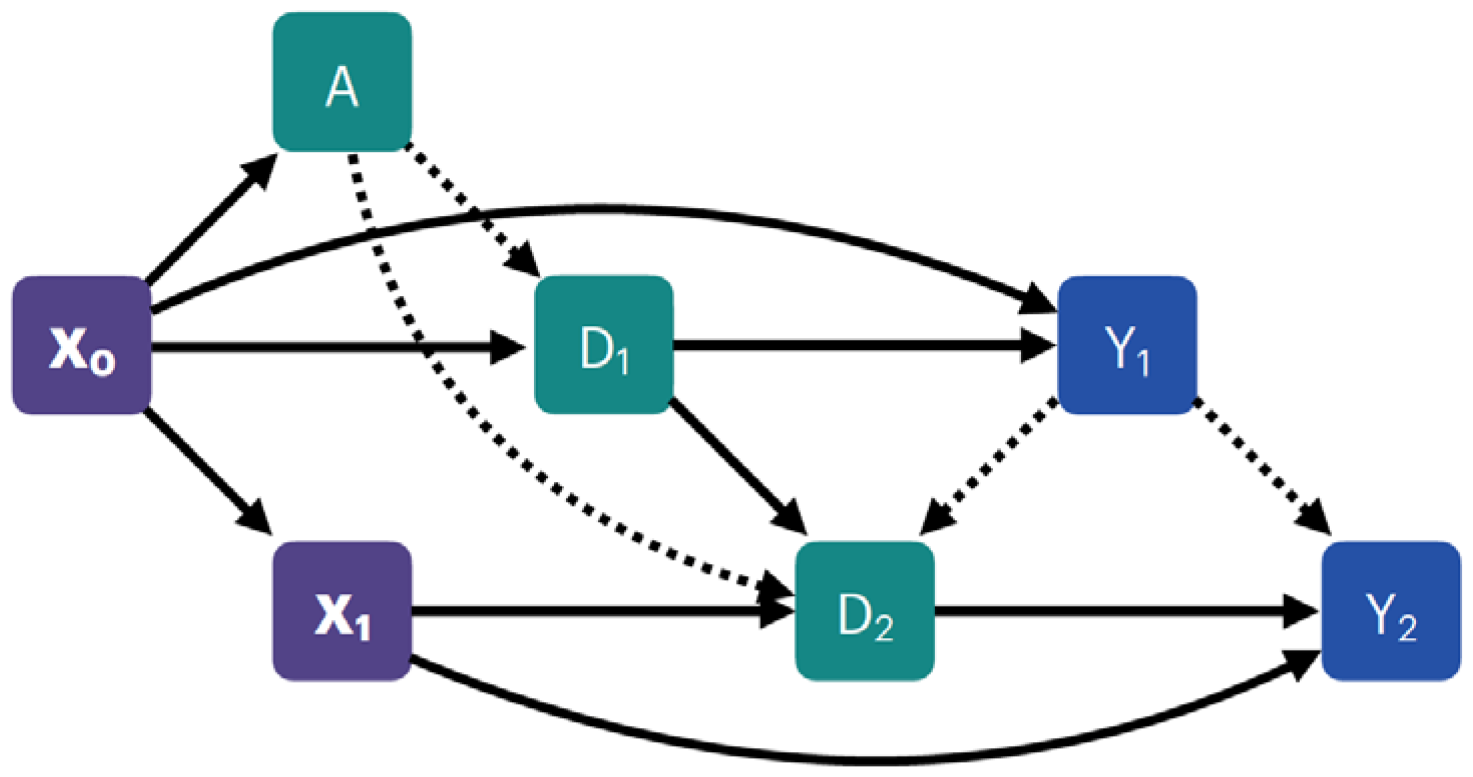
DAG representing the relationships among the exposures (*A, D*_1_, *D*_2_), the outcomes (*Y*_1_, *Y*_2_), and the covariates (***X***_0_,***X***_1_). Dashed lines indicate deterministic relationships.

#### A Analysis

We first evaluate the effect of access (*A*) on risk of HIV within 1 year (*Y*_2_) by considering the causal relative risk (RR)

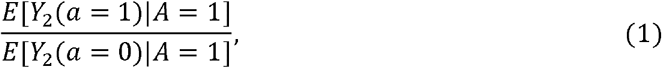

where *Y*_2_ (*a*) is the potential outcome of *Y*_2_ under exposure *A* = *a*, i.e., the outcome that would have been observed had *A* = *a* [27]. This is the causal RR of HIV infection in one year comparing PrEP access versus no access in the HPTN 073 population, that is, conditioned on *A* = 1 [28].

One approach to estimating (1) is IPW, which uses weights to balance the distribution of covariates between HPTN 061 and HPTN 073, thereby mimicking a randomized trial in which the only (systematic) difference between the exposure groups is the exposure itself [29]. While the ideal would be to make the HPTN 061 sample similar to the HPTN 073 sample on all covariates, we can only achieve similarity for the measured covariates [30]. Here, the goal of weighting was to make the distribution of covariates in the HPTN 061 sample similar to that in the HPTN 073 sample because the HPTN 073 population was our target of inference. In other words, we used weights to obtain a sample from a “pseudopopulation” in which the distribution of the measured baseline covariates in both the exposed (*A* = 1) and the unexposed (*A* = 0) reflects the distribution of the covariates in the HPTN 073 population [20, 28, 31, 32], removing the potential for confounding by these variables [20, 32].

We can estimate (1) using the HPTN 061 and HPTN 073 samples with weights of the form [28, 33]

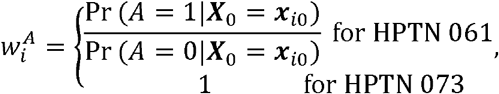

*i* = 1, …, *n*. using 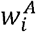, (1) can be estimated by fitting a weighted log-linear model of the form

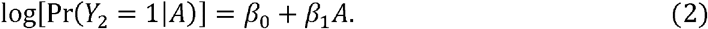

This approach requires assuming conditional exchangeability, i.e., no unmeasured confounding, and positivity, that is, a positive probability of *A* = 0 [34, 35]. These assumptions are formally stated and described in greater detail in the Supplement, along with a few other standard assumptions and the derivation of the weights 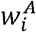.

To obtain 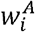, an estimate of Pr (*A* = 1 | ***X***_0_) is needed. As we sought weights that would make the HPTN 061 sample similar to the HPTN 073 sample in terms of the distribution of the covariates, we used the covariate-balancing propensity score (CBPS), which estimates Pr (*A* = 1 | ***X***_0_) by modeling treatment assignment while optimizing (mean) covariate balance [36, 37]. Our implementation relied on the CBPS package in R [38, 39].

#### D Analysis

The causal effect of PrEP initiation and adherence (*D*), a time-varying variable, can be written as

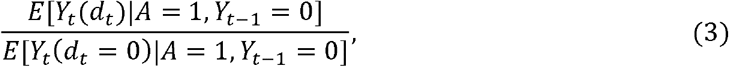

where *t* = 1,2, *d*_*t*_ *=*1,2. Here, the potential outcome *Y*_*t*_ (*d*_*t*_) is the outcome an individual would have experienced (in this case HIV infection by visit) had they received exposure *d*_*t*_ during the prior time interval. The estimand in (3) yields the RR of HIV infection during the next six months for (i) individuals who had initiated PrEP but were not adherent at time *t* vs. those who had not yet initiated PrEP(*d*_*t*_ *=*1 vs. 0) and (ii) individuals who were adherent to PrEP at time *t* vs. those who had not yet initiated PrEP (*d*_*t*_ = 2 vs.0) [40]. We conditioned on *Y*_*t* −1_ = 0 because individuals who already tested positive for HIV are no longer in the risk set and on *A* = 1because the HPTN 073 population was our target of inference.

As we wanted to (i) use data from HPTN 073, a non-randomized study, and (ii) incorporate data from HPTN 061, a single-arm study in which PrEP was not available, two sets of weights were required. The first set of weights was used to make the distribution of the covariates in the HPTN 061 population similar to that in the HPTN 073 population:

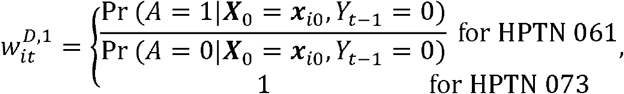

*t* = 1,2, *i* = 1,…,*n*.The second set of weights was used to achieve balance in the distribution of measured covariates for *D*_*t*_ = 0,1, and 2 in the HPTN 073 sample:

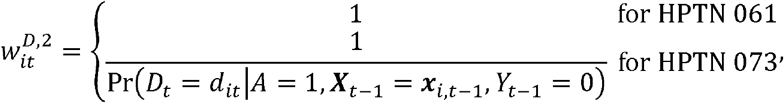

where *i* = 1,…,*n,t* = 1,2. The final weights were obtained by multiplying 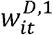 and 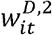 yielding

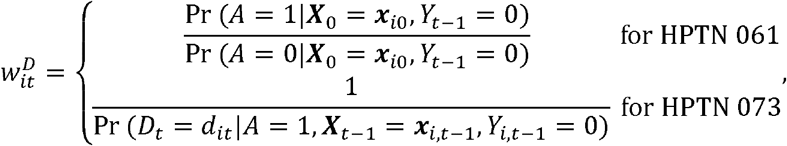

where and *i* = 1,…,*n,t* = 1,2, and *d*_*t*_ = 0,1,2. More details on these weights and their estimation with CBPS are given in the Supplement.

Using the weights 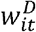, (3) was estimated by fitting a weighted log-linear model [41] among individuals who have not yet tested positive for HIV, similar to a pooled logistic regression model [32, 42] (more details in the Supplement):

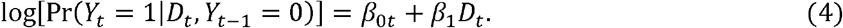

This approach requires an assumption of conditional exchangeability for both *A* and *D*, i.e., no unmeasured confounding, and positivity, that is, a positive probability of *A* = 0 and *D* ∈ {0,1,2} [34, 43]. These assumptions are formally stated and described in greater detail in the Supplement, along with a few other standard assumptions and the derivation of the weights 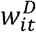.

### Covariate Selection, Missing Data, and Quantifying Uncertainty

For inverse probability weights, covariates related to both the outcome and the exposure should be included [44], though the inclusion of covariates only related to the outcome may increase efficiency without incurring bias [45-47]. Furthermore, including too many covariates can increase the likelihood of positivity violations [48-50]. Thus, we focused only on covariates that are believed or known to be causes of HIV [47, 51].

The risk of infection with HIV in any population, including Black MSM in the US, is a function of an individual’s sexual network and the frequency of condomless sexual encounters, and may be augmented by cofactors such as concomitant STIs. These features correspond to known risk factors for HIV [21, 52] and are plausible causal agents given what is known about HIV transmission in this population. Thus, we chose to include region (categorial), age (continuous), number of partners (0-1, 2-3, 4 or more; ordinal), sexual orientation (gay vs. other), condomless receptive anal intercourse (URAI; binary), and the presence of an STI (binary), each of which captures a dimension of the above features. Number of partners, URAI, and STI were time-varying covariates.

Missing data were largely accommodated through multiple imputation. To quantify uncertainty, we generated 95% confidence intervals (CIs) using the bootstrap, as recommended in this setting [53-55]. See the Supplement for additional details on both missing data and interval estimation.

## RESULTS

### Effect of PrEP Initiation and Adherence

Table 1 summarizes the key characteristics of the HPTN 061 and HPTN 073 samples. There were 1,134 individuals in the HPTN 061 sample (after removing those ineligible for HPTN 073) and 226 individuals in the HPTN 073 sample. In HPTN 073, more than three-quarters of participants initiated PrEP in the first 6 months, with few new initiators in the second interval. Nearly 44% of all participants (57.2% of PrEP initiators) were adherent to PrEP in the first six months; this decreased to 36.3% of all participants (46.1% of PrEP initiators) in the second six in the second six months. In HPTN 061, 18 individuals tested positive for HIV by the six-month visit (*t* = 1); by the twelve-month visit (*t* = 2), a total of 28 individuals had tested positive (i.e., there were 10 new seroconversions between the six- and twelve-month visits). In HPTN 073, 5 individuals tested positive by the six-month visit (*t* = 1) and a total of 8 tested positive by the twelve-month visit (*t* = 1). The degree of missingness of HIV status was considerably higher in HPTN 061 compared to HPTN 073, particularly at the twelve-month time point. There were several important differences in the relevant covariates between HPTN 061 and HPTN 073 and in the extent of missingness. For both studies, the extent of missing covariate values was low at baseline, though this increased substantially at the 6-month time point (*t* = 1).

**Table 1:**
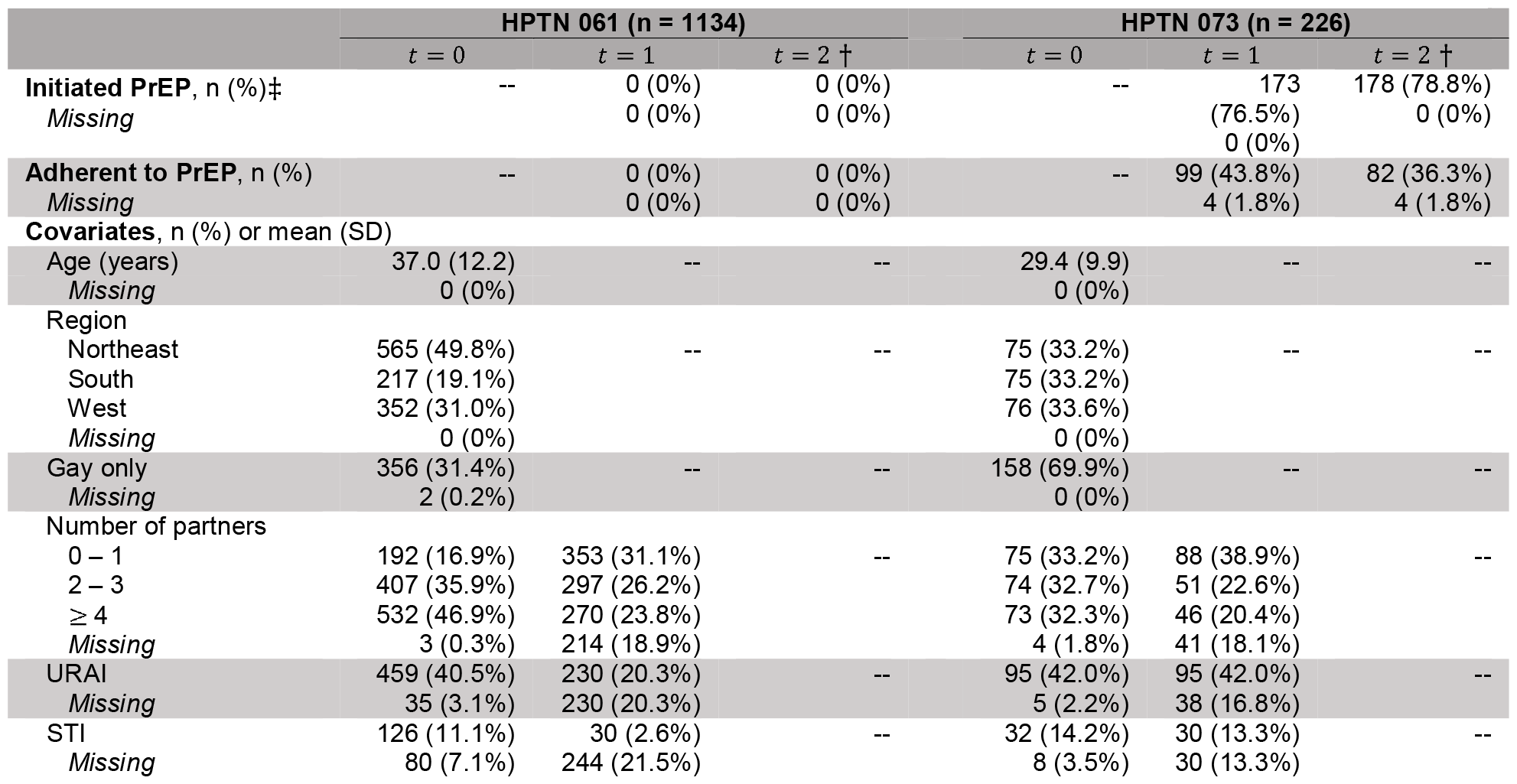
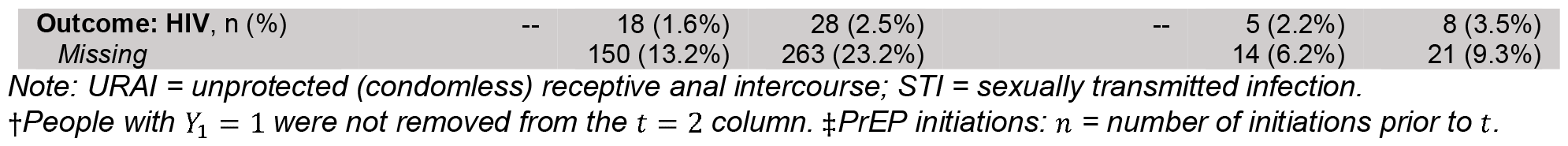
Cohort summaries for the HPTN 061 sample and the HPTN 073 sample.

Based on the weighted analysis, we estimated a 48% reduction in risk of HIV in 1 year due to having access to PrEP (0.520, 95% CI for the RR: 0.213, 1.217) (Table 2). PrEP initiation (without adherence) during a six-month interval resulted in an estimated 52% reduction in risk of HIV at the end of the interval compared to individuals who did not initiate PrEP (95% CI for the RR: (0.122, 0.893)) and PrEP adherence during a six-month interval resulted in an estimated 77% reduction in risk of HIV at the end of the interval compared to individuals who did not initiate PrEP (95% CI for the RR: 0.015, 0.798). Thus, we observed substantial reductions in the estimated risk of HIV due to PrEP access, initiation, and adherence, though only the latter two were significant at the 0.05 level. Importantly, these estimates pertain to the HPTN 073 population.

**Table 2:**
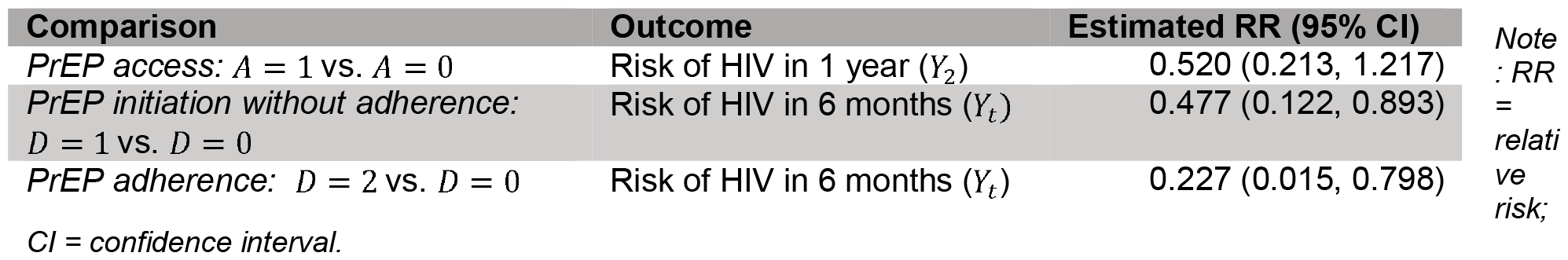
Effect estimates for PrEP access, initiation, and adherence.

### Diagnostics & Sensitivity Analyses

We used established diagnostics to evaluate balance of measured covariates and the presence of extreme probabilities (i.e., violations of the positivity assumptions); the analyses are described and the results presented in the Supplement. Briefly, good covariate balance was achieved and, while extreme probabilities were found, these were generally only in the bootstrap samples, yielding wider confidence intervals. Finally, we used established methods to evaluate the impact of unmeasured confounders [19, 41, 56, 57]. Overall, we found that the causal effects of PrEP access, initiation, and adherence on HIV risk were attenuated in the presence of an unmeasured confounder with a strong negative association with HIV risk.

## DISCUSSION

We have estimated the causal effects of PrEP access, initiation (without adherence), and adherence in Black MSM in the US based on a careful analysis combining data from two non-randomized studies, HPTN 061 and HPTN 073. Using a weighting approach, we found that in the HPTN 073 population, access to PrEP reduced risk of HIV by an estimated 48% and, compared to Black MSM who did not initiate PrEP, PrEP initiation reduced HIV risk by an estimated 52% and PrEP adherence reduced HIV risk by an estimated 77%, though only the latter two estimates were statistically significant. These large estimated reductions in risk of HIV are not surprising, given the large protective effects of PrEP observed in other populations [11, 12, 14, 17, 58]. Our results are of considerable public health importance as they provide estimates of the effect of PrEP specific to a population that bears a disproportionate burden of the HIV epidemic in the US. These findings may motivate some men to initiate and adhere to PrEP, substantially reducing their risk of HIV infection. While our analysis was well-motivated and made judicious use of existing data, alternatives exist and are assessed in the Supplement.

Importantly, beyond the specific goal of estimating the causal effect of PrEP in Black MSM in the US, our approach provides a framework for combining data from multiple non-randomized studies to estimate causal effects. This framework may be useful for estimating the effect of PrEP in other populations and, more broadly, for estimating causal effects of other exposures using existing datasets. Such strategies are of particular value in the context of large networks, like HPTN, which offer the opportunity to utilize data from multiple studies to answer causal questions without a randomized trial. Furthermore, when datasets like these exist, there is an ethical obligation to utilize the data as fully as possible, using robust and rigorous methods. Such stewardship is particularly necessary when the data come from minoritized communities, as these communities are frequently called upon to participate in research and may be overburdened or wary of participation altogether due to past encounters with the medical community.

### Strengths

Our analysis involved data from two carefully conducted observational studies, including one fairly large study. Since both studies were under the umbrella of HPTN and conducted reasonably close together in time, data collection (variables measured, definitions used, and assessment mechanisms) and recruitment and retention strategies were rigorous and similar between the two studies, making their combination highly compelling. While some challenges related to combining the data from HPTN 061 and HPTN 073 persist (as discussed below), this analysis would not have been possible had we been limited to only one of these datasets given the size of HPTN 073 and lack of PrEP in HPTN 061. Importantly, individuals who enroll in observational studies, like HPTN 061 and HPTN 073, may be more representative of the general population than those who would enroll in a randomized trial, enhancing the generalizability of the results.

We utilized a rigorous and robust approach to effect estimation, quantification of uncertainty, and evaluation of the plausibility of key assumptions. In so doing, we have provided a framework that can be used more generally to estimate causal effects using data from multiple non-randomized studies. Even when a randomized controlled trial is ethical and feasible, if data from non-randomized studies exists, it may be advisable to employ a framework like ours to either provide motivation for a randomized trial or, if the data are from well-conducted studies, to obviate the need for a trial altogether, preserving financial resources, reducing burden on would-be participants, and allowing the expedient estimation of causal effects.

### Limitations

The HPTN 073 sample may not be representative of the HPTN 073 population [24, 59] and the HPTN 073 population may differ from our target population, Black MSM in the US [24, 60]. Generalizing our results to the population of Black MSM in the US requires careful consideration of selection bias [61] and, relatedly, the sampling schemes, enrollment processes, and eligibility criteria used. While some degree of generalizability may be lost, this will only be a concern insofar as there is effect modification and/or unmeasured confounding [62]. Residual imbalances between HPTN 061 and HPTN 073 may persist owing to imperfect overlap in the variables measured and the possibility of differences in the distribution of effect modifiers not included in our weight models [24]. Importantly, although the use of study indicator as a proxy for PrEP access implicitly assumes no effect of study apart from PrEP access, this is likely to be reasonable given the similarity between the two studies.

Differences in the frequency of data collection between the two studies and use of self-report for several covariates may have resulted in measurement error (see the Supplement for a discussion of this issue) [21, 63]. Additionally, the reliance on assessments of PrEP initiation, PrEP adherence, and HIV status every 6 months (as opposed to a shorter time interval) may have led to measurement error and some degree of reverse causation that could have biased results if individuals obtained off-study HIV tests. Future analyses based on data with more frequent assessments would be informative.

### Conclusions

We have estimated the causal effects of PrEP access, initiation, and adherence in a key population in the US HIV epidemic, Black MSM, finding large reductions in risk, particularly for initiation and adherence. More broadly, we have created a rigorous analytic framework for combining data from multiple studies to estimate causal effects. This framework may be generally useful when seeking to estimate causal effects using several data sources, particularly when a randomized control trial is unethical or infeasible.

## DATA AVAILABILITY

Data and code to run this analysis are available at: https://github.com/meisnera/BMSM_PrEP.

## ACKNOWLEDGEMENTS

We acknowledge funding from NIH U01 HL146242 and NIH P30 MH123248. Overall support for the HIV Prevention Trials Network (HPTN) is provided by the NIH under Award Numbers UM1AI068619 (HPTN Leadership and Operations Center), UM1AI068617 (HPTN Statistical and Data Management Center), and UM1AI068613 (HPTN Laboratory Center). Additional support for this study was provided by UM1AI069412. We extend our thanks the HPTN 061 and HPTN 073 study teams and study participants.

## Notes

### Competing Interest Statement

The authors have declared no competing interest.

### Clinical Trial

NCT00951249, NCT01808352

### Author Declarations

The studies (HPTN 061 and HPTN 073) were reviewed and approved by the IRBs of each study site and all participants provided written informed consent, including for the data analysis presented in this work.

### Summary of Updates

Paper length reduced; materials moved to the supplement.

## REFERENCES

1. HIV.gov. Data & Trends: U.S. Statistics. 2022 [cited 2023 August 15]; Available from: https://www.hiv.gov/hiv-basics/overview/data-and-trends/statistics/.

2. US Census Quick Facts. 2020 [cited 2023 September 14]; Available from: https://www.census.gov/quickfacts/fact/table/US/IPE120221.

3. Brewer, R.A., et al., Exploring the relationship between incarceration and HIV among black men who have sex with men in the United States. JAIDS: Journal of acquired immune deficiency syndromes 2014. 65(2): p. 218.

4. Fields, E.L., et al., “I always felt I had to prove my manhood”: Homosexuality, masculinity, gender role strain, and HIV risk among young Black men who have sex with men. American Journal of Public Health, 2015. 105(1): p. 122–131.

5. Levy, M.E., et al., Understanding structural barriers to accessing HIV testing and prevention services among black men who have sex with men (BMSM) in the United States. AIDS and Behavior, 2014. 18: p. 972–996.

6. Malebranche, D.J., et al., Race and sexual identity: perceptions about medical culture and healthcare among Black men who have sex with men. Journal of the National Medical Association, 2004. 96(1): p. 97.

7. Mayer, K.H., et al., Concomitant socioeconomic, behavioral, and biological factors associated with the disproportionate HIV infection burden among Black men who have sex with men in 6 US cities. PLOS One, 2014. 9(1): p. e87298.

8. Mena, L., R.A. Crosby, and A. Geter, A novel measure of poverty and its association with elevated sexual risk behavior among young Black MSM. International Journal of STD & AIDS, 2017. 28(6): p. 602–607.

9. Millett, G.A., et al., Comparisons of disparities and risks of HIV infection in black and other men who have sex with men in Canada, UK, and USA: a meta-analysis. The Lancet, 2012. 380(9839): p. 341–348.

10. Nelson, L.E., et al., Economic, legal, and social hardships associated with HIV risk among black men who have sex with men in six US cities. Journal of Urban Health, 2016. 93: p. 170–188.

11. Baeten, J.M., et al., Antiretroviral prophylaxis for HIV prevention in heterosexual men and women. New England Journal of Medicine, 2012. 367(5): p. 399–410.

12. Choopanya, K., et al., Antiretroviral prophylaxis for HIV infection in injecting drug users in Bangkok, Thailand (the Bangkok Tenofovir Study): a randomised, double-blind, placebo-controlled phase 3 trial. The Lancet, 2013. 381(9883): p. 2083–2090.

13. Donnell, D., et al., HIV protective efficacy and correlates of tenofovir blood concentrations in a clinical trial of PrEP for HIV prevention. JAIDS: Journal of Acquired Immune Deficiency Syndromes, 2014. 66(3): p. 340.

14. Grant, R.M., et al., Preexposure chemoprophylaxis for HIV prevention in men who have sex with men. New England Journal of Medicine, 2010. 363(27): p. 2587–2599.

15. Marrazzo, J.M., et al., Tenofovir-based preexposure prophylaxis for HIV infection among African women. New England Journal of Medicine, 2015. 372(6): p. 509–518.

16. Murnane, P.M., et al., Efficacy of pre-exposure prophylaxis for HIV-1 prevention among high risk heterosexuals: subgroup analyses from the Partners PrEP Study. AIDS, 2013. 27(13).

17. Thigpen, M.C., et al., Antiretroviral preexposure prophylaxis for heterosexual HIV transmission in Botswana. New England Journal of Medicine, 2012. 367(5): p. 423–434.

18. Van Damme, L., et al., Preexposure prophylaxis for HIV infection among African women. New England Journal of Medicine, 2012. 367(5): p. 411–422.

19. Robins, J.M., Association, causation, and marginal structural models. Synthese, 1999. 121(1/2): p. 151–179.

20. Robins, J.M., M.A. Hernan, and B. Brumback, Marginal structural models and causal inference in epidemiology. Epidemiology, 2000: p. 550–560.

21. Koblin, B.A., et al., Correlates of HIV acquisition in a cohort of Black men who have sex with men in the United States: HIV prevention trials network (HPTN) 061. PLOS One 2013. 8(7): p. e70413.

22. Wheeler, D.P., et al., Pre-exposure prophylaxis initiation and adherence among Black men who have sex with men (MSM) in three US cities: results from the HPTN 073 study. Journal of the International AIDS Society, 2019. 22(2): p. e25223.

23. Hendrix, C.W., et al., Dose frequency ranging pharmacokinetic study of tenofovir-emtricitabine after directly observed dosing in healthy volunteers to establish adherence benchmarks (HPTN 066). AIDS Research and Human Retroviruses, 2016. 32(1): p. 32–43.

24. Degtiar, I. and S. Rose, A review of generalizability and transportability. Annual Review of Statistics and Its Application, 2023. 10: p. 501–524.

25. Levy, M.E., et al., A longitudinal analysis of treatment optimism and HIV acquisition and transmission risk behaviors among black men who have sex with men in HPTN 061. AIDS and Behavior, 2017. 21: p. 2958–2972.

26. Whitfield, D.L., et al., Risk compensation in HIV PrEP adherence among Black men who have sex with men in HPTN 073 study. AIDS Care, 2021. 33(5): p. 633–638.

27. Hernán, M. and J. Robins, Causal Inference: What If? 2020, Boca Raton: Chapman & Hall/CRC.

28. Sato, T. and Y. Matsuyama, Marginal structural models as a tool for standardization. Epidemiology, 2003: p. 680–686.

29. Hernán, M.A. and J.M. Robins, Using big data to emulate a target trial when a randomized trial is not available. American Journal of Epidemiology, 2016. 183(8): p. 758–764.

30. Platt, R.W., J.A.C. Delaney, and S. Suissa, The positivity assumption and marginal structural models: the example of warfarin use and risk of bleeding. European Journal of Epidemiology, 2012. 27: p. 77–83.

31. Austin, P.C., An introduction to propensity score methods for reducing the effects of confounding in observational studies. Multivariate Behavioral Research, 2011. 46(3): p. 399–424.

32. Hernán, M.Á., B. Brumback, and J.M. Robins, Marginal structural models to estimate the causal effect of zidovudine on the survival of HIV-positive men. Epidemiology, 2000: p. 561–570.

33. Li, F., K.L. Morgan, and A.M. Zaslavsky, Balancing covariates via propensity score weighting. Journal of the American Statistical Association, 2018. 113(521): p. 390–400.

34. Imbens, G.W., The role of the propensity score in estimating dose-response functions. Biometrika, 2000. 87(3): p. 706–710.

35. Westreich, D., et al., Imputation approaches for potential outcomes in causal inference. International Journal of Epidemiology, 2015. 44(5): p. 1731–1737.

36. Imai, K. and M. Ratkovic, Robust estimation of inverse probability weights for marginal structural models. Journal of the American Statistical Association, 2015. 110(511): p. 1013–1023.

37. Imai, K. and M. Ratkovic, Covariate balancing propensity score. Journal of the Royal Statistical Society Series B: Statistical Methodology, 2014. 76(1): p. 243–263.

38. Fong C R.M., Imai K CBPS: Covariate Balancing Propensity Score. 2022. p. R package.

39. R Core Team R: A language and environment for statistical computing 2023, R Foundation for Statistical Computing: Vienna, Austria.

40. Ko, H., J.W. Hogan, and K.H. Mayer, Estimating causal treatment effects from longitudinal HIV natural history studies using marginal structural models. Biometrics, 2003. 59(1): p. 152–162.

41. Chiba, Y., Sensitivity analysis of unmeasured confounding for the causal risk ratio by applying marginal structural models. Communications in Statistics - Theory and Methods, 2009. 39(1): p. 65–76.

42. Ngwa, J.S., et al., A comparison of time dependent Cox regression, pooled logistic regression and cross sectional pooling with simulations and an application to the Framingham Heart Study. BMC Medical Research Methodology, 2016. 16: p. 1–12.

43. Ertefaie, A. and D.A. Stephens, Comparing approaches to causal inference for longitudinal data: inverse probability weighting versus propensity scores. The International Journal of Biostatistics, 2010. 6(2).

44. Fewell, Z., et al., Controlling for time-dependent confounding using marginal structural models. The Stata Journal, 2004. 4(4): p. 402–420.

45. Austin, P.C., P. Grootendorst, and G.M. Anderson, A comparison of the ability of different propensity score models to balance measured variables between treated and untreated subjects: a Monte Carlo study. Statistics in Medicine, 2007. 26(4): p. 734–753.

46. Brookhart, M.A., et al., Variable selection for propensity score models. American Journal of Epidemiology, 2006. 163(12): p. 1149–1156.

47. Shiba, K. and T. Kawahara, Using propensity scores for causal inference: pitfalls and tips. Journal of Epidemiology, 2021. 31(8): p. 457–463.

48. Crowson, C.S., et al., The Basics of Propensity Scoring and Marginal Structural Models 2013: Mayo Clinic.

49. McCulloch, C.E., observational studies, time-dependent confounding, and marginal structural models. Arthritis & Rheumatology, 2015. 67(3): p. 609–611.

50. Xiao, Y., M. Abrahamowicz, and E.E. Moodie, Accuracy of conventional and marginal structural Cox model estimators: a simulation study. The International Journal of Biostatistics, 2010. 6(2).

51. Hernán, M.A., B.A. Brumback, and J.M. Robins, Estimating the causal effect of zidovudine on CD4 count with a marginal structural model for repeated measures. Statistics in Medicine, 2002. 21(12): p. 1689–1709.

52. Garofalo, R., et al., Incidence of HIV infection and sexually transmitted infections and related risk factors among very young men who have sex with men. JAIDS Journal of Acquired Immune Deficiency Syndromes, 2016. 72(1): p. 79.

53. Cole, S.R., et al., Effect of highly active antiretroviral therapy on time to acquired immunodeficiency syndrome or death using marginal structural models. American Journal of Epidemiology, 2003. 158(7): p. 687–694.

54. Reifeis, S.A. and M.G. Hudgens, On variance of the treatment effect in the treated when estimated by inverse probability weighting. American Journal of Epidemiology, 2022. 191(6): p. 1092–1097.

55. Xiao, Y., E.E. Moodie, and M. Abrahamowicz, Comparison of approaches to weight truncation for marginal structural Cox models. Epidemiologic Methods, 2013. 2(1): p. 1–20.

56. Brumback, B.A., et al., Sensitivity analyses for unmeasured confounding assuming a marginal structural model for repeated measures. Statistics in Medicine, 2004. 23(5): p. 749–767.

57. Klungsøyr, O., et al., Sensitivity analysis for unmeasured confounding in a marginal structural Cox proportional hazards model. Lifetime Data Analysis, 2009. 15: p. 278–294.

58. Molina, J.-M., et al., On-demand preexposure prophylaxis in men at high risk for HIV-1 infection. New England Journal of Medicine, 2015. 373(23): p. 2237–2246.

59. Naimi, A.I., S.R. Cole, and E.H. Kennedy, An introduction to g methods. International Journal of Epidemiology, 2017. 46(2): p. 756–762.

60. Olsen, R.B., et al., External validity in policy evaluations that choose sites purposively. Journal of Policy Analysis and Management, 2013. 32(1): p. 107–121.

61. Haneuse, S., et al., Adjustment for selection bias in observational studies with application to the analysis of autopsy data. Neuroepidemiology, 2009. 32(3): p. 229–239.

62. Choi, J., O.M. Dekkers, and S. le Cessie, A comparison of different methods to handle missing data in the context of propensity score analysis. European Journal of Epidemiology, 2019. 34: p. 23–36.

63. Luehring-Jones, P., et al., Pre-exposure prophylaxis (PrEP) use is associated with health risk behaviors among moderate-and heavy-drinking MSM. AIDS Education and Prevention, 2019. 31(5): p. 452–462.

