## Supplement for "Estimating the Effect of PrEP in Black Men Who Have Sex with Men: A Framework to Utilize Data from Multiple Non-Randomized Studies to Estimate Causal Effects"

**SUPPLEMENTARY MATERIALS**

*Details of HPTN 061*

Men were eligible to participate in HPTN 061 if they: identified as a man or male at birth and as Black, African American, Caribbean Black, or multiethnic Black; were at least 18 years old; reported $\geq$ 1 instance of unprotected anal intercourse with a man in the prior six months; and resided in the metropolitan area and did not plan to move [1]. Men were followed up with visits 6 months (80.3% completion) and 12 months (74.9% completion) after enrollment; at these visits, HIV status was determined for 86.5% of participants and 76.5% of participants, respectively [1]. HPTN 061 was registered on ClinicalTrials.gov (NCT00951249).

*Details of HPTN 073*

Men were eligible to participate in HPTN 073 if they: self-identified as Black; were at least 18 years old; met safety criteria for PrEP eligibility; had not used antiretroviral drugs, including PrEP or post-exposure prophylaxis (PEP), in the previous 60 days; and reported at least one of the following in the previous 6 months: unprotected anal intercourse with a male partner, any anal intercourse with more than 3 male sex partners, exchange of anal sex with a male partner for money or goods, anal sex with a male partner while using drugs or alcohol, or diagnosis of a sexually transmitted infection (STI) and sex with a male partner [2]. Individuals were not eligible to participate if they had hepatitis B, a prior history of certain medical procedures, and/or received prohibited medications [2]. Retention was quite high with 92% of enrolled participants completing the week 52 follow-up visit. Adherence was defined as at least 4 doses of PrEP per week, i.e., the individual had either (a) tenofovir (TFV) levels of at least 4.2 ng/mL and FTC levels of at least 4.6 ng/mL in plasma or (b) TFV diphosphate levels of at least 9.9 fmol/10^6^ and FTC triphosphate levels of at least 0.4 fmol/10^6^ in peripheral blood mononuclear cells (PMBCs) [3]. All individuals enrolled in HPTN 073 had access to the C4 program, a light-touch support program [22], though it is reasonable to assume that C4 does not have a direct effect of HIV risk. HPTN 073 was registered on ClinicalTrials.gov (NCT01808352).

*Details on Reconciling HPTN 061 and HPTN 073*

Some differences in inclusion criteria in HPTN 061 and HPTN 073 led to entire groups being excluded from one of the studies but not the other. For instance, HPTN 073 did not allow transgender males to enroll; thus, we excluded transgender males from HPTN 061 (n = 20) before combining the two datasets. HPTN 073 also required participants to have had sex with a man (who was born male) in the previous 6 months, while HPTN 061 required sex with a man (gender assigned at birth not specified) in the same time period. Thus, we further excluded individuals in HPTN 061 who reported only having transgender male partners in the past six months at baseline (n = 8). Discrepancies in inclusion criteria between the two studies were expected to lead to differences in the distribution of some covariates between the two studies; such imbalances were typically accommodated by weighting [4, 5]. Remaining discrepancies include the fact that HPTN 073 excluded individuals who did not meet the clinical safety criteria for PrEP eligibility [2]. However, since HPTN 061 took place in the pre-PrEP era, data related to PrEP eligibility criteria were not collected in HPTN 061, so a similar exclusion was not possible in HPTN 061. Additionally, individuals who had used antiretroviral drugs, including PrEP or post-exposure prophylaxis (PEP), in the previous 60 days were excluded from participating in HPTN 073. While a very small number of HPTN 061 participants reported *ever* using PrEP/PEP, the exclusion in HPTN 073 only related to PrEP/PEP use in the last 60 days, so we assumed that almost no one in HPTN 061 used PEP in the last 60 days. Some other clinical exclusions in HPTN 073 related to hepatitis B infection, prior history of certain medical procedures, and receipt of prohibited medications could not be extended to HPTN 061 as the relevant data were not collected. However, the impact of these discrepancies was expected to be minimal.

Another challenge is that in HPTN 061, when participants were asked about sexual behaviors (e.g., number of partners, HIV status of partners, etc.), the questions asked about all partners who were men, while in HPTN 073, these questions asked only about partners who were born male. In order to minimize loss of information, we conceptualized these questions as pertaining to partners who were born male or were transgender male, as in HPTN 061. In HPTN 073, for participants with only transgender male partners, the values of these sexual behavior covariates were imputed. Given that the participants in HPTN 061 tended to have far more partners who were born male than transgender male partners, the participants’ responses to these questions largely reflect their experiences with partners who were born male, making them likely very similar to the responses that would have been obtained had they been enrolled in HPTN 073. Further, as there were very few men in HPTN 073 with transgender male partners (and only a handful with *only* transgender male partners), this was a reasonable approach and was expected to minimize both measurement error and loss of information.

Relatedly, data collection was more frequent in HPTN 073 (every three months) than in HPTN 061 (every six months). One consequence is that, at each visit, the questions asked of participants in HPTN 073 involved a three-month lookback, while the questions asked of HPTN 061 participants pertained to the previous six months. Since we only used the measurements from the 6-month visit to estimate weights, some information from HPTN 073 regarding time-varying covariates was lost. Consequently, there may have been some measurement error arising from the utilization of covariate information collected at the 6-month visits in HPTN 073 to reflect the previous six months, when in fact it only captures the previous three months. We attempted to mitigate this source of measurement error by using the data collected at the 3-month visit in the case of binary time-varying covariates. In particular, for these binary (yes/no) time-varying covariates, if a participant responded “yes” at either the 3- or 6-month visit this was counted as a “yes”. Most of the time-varying covariates used to estimate the weights were binary, as were many of the variables used in imputation (which also utilized covariates measured at the 12-month visit; the procedure described above was deployed to incorporate information collected at the 9-month visit in HPTN 073 in the same way as the covariates measured at the 3-month visit). Thus, this source of measurement error is not expected to have a meaningful impact.

*Population Definitions*

Given that the estimates and inference resulting from our analysis will depend to some degree on the distribution of covariates when there is effect modification [6, 7] and/or unmeasured confounding, differences in covariate distributions may lead to issues with interpretability and/or generalizability if the relevant populations are not well-defined [8]. Thus, careful definitions of the target populations, study populations, and study samples (given in the main paper) are needed. Importantly, study populations are defined by enrollment processes and eligibility criteria, e.g., all Black MSM living in certain US cities at the time of the study and satisfying all eligibility criteria; study samples are then (randomly) drawn from these populations [7].

$A$ *Analysis Assumptions*

In order to use the inverse probability weights $w_{i}^{A}$ to estimate $\frac{E[Y_{2}(a=1)|A=1]}{E[Y_{2}(a=0)|A=1]}$, some key assumptions are required [9, 10]:

(a) Conditional exchangeability: $Y_{2}\left( a=0 \right)\perp A | \boldsymbol{X}_{0}$ (assumption A1)
(b) Positivity: $\Pr\left( A=0 | \boldsymbol{X}_{0}=\boldsymbol{x}_{0} \right)>0 \forall\boldsymbol{x}_{0}$ (assumption A2)

The assumption of conditional exchangeability (A1) means that conditional on the covariates $\boldsymbol{X}_{0}$, the distribution of $Y_{2}(a=0)$ is the same in the HPTN 061 population and in the HPTN 073 population [11, 12]. This allows us to use the observed outcomes for $A=0$ (i.e., $Y_{2}$ in the HPTN 061 population) in place of the potential outcomes $Y_{2}(a=0)$ for $A=1$, permitting estimation of $\frac{E[Y_{2}(a=1)|A=1]}{E[Y_{2}(a=0)|A=1]}$ [13]. The idea behind (A1) is that it allows us to use the weights $w_{i}^{A}$ to understand what would have happened to the HPTN 073 population (in terms of HIV risk) had there been no access to PrEP [12]. Assumption (A1) can also be thought of as an assumption of no unmeasured confounding of the association between PrEP access and HIV risk; essentially, we allowed for confounding by $\boldsymbol{X}_{0}$ but not by any other factors (see DAG in Figure 1) [12, 14, 15]. The idea is that the weights aimed to provide balance on the measured covariates, and (A1) implies unmeasured covariates were not relevant [9]. We do not need to make a similar assumption for $Y_{2}(a=1)$ because these potential outcomes are observed in the HPTN 073 population, the target of inference [13].

The assumption of positivity (A2) makes $Y_{2}(a=0)$ realizable for all participants and thus allows us to estimate $E(Y_{2}(a=0)|A=1)$ [11, 13]; note that $E(Y_{2}(a=1)|A=1)$ can be estimated using the HPTN 073 sample, provided consistency (discussed below) holds. As with (A1), we did not need to make an assumption about $\Pr\left( A=1 | \boldsymbol{X}_{0} \right)$ because (in theory) it would be permissible to include some HPTN 061 participants who were ineligible for HPTN 073 though they would not contribute to estimation [13] (though in reality the presence of such individuals would lead to difficulties in estimating $\Pr\left( A=1 | \boldsymbol{X}_{0} \right))$. The assumption (A2) essentially requires that all members of the HPTN 073 population are represented by individuals in the HPTN 061 population [7]. Near-violations of (A2) may also present difficulties, as this could mean that massive weights are given to some observations, which would then have an outsized influence on the results, potentially leading to bias, instability, and increased uncertainty [6, 16-19]. Importantly, issues with positivity (violations of (A2)) are not anticipated as the HPTN 073 and HPTN 061 populations are quite similar.

While (A1) and (A2) were the key assumptions, we also assumed consistency ($A=a \Rightarrow Y_{2}=Y_{2}(a)$ for a particular subject) [10, 16] and the stable unit treatment value assumption (SUTVA), which supposes that there were not multiple versions of exposure (i.e., the exposures A = 0 and A = 1 are well-defined and homogeneous) and that each individual’s potential outcomes were independent of the exposure status of other individuals, i.e., there was no interference [20]. Consistency generally holds if the recorded exposure (in this case, A) reflects the exposure the participant actually experienced [21].

$D$ *Analysis Weights, Model, and Assumptions*

The weights $w_{it}^{D,1}$are quite similar to $w_{i}^{A},$ but are time-varying since they condition on $Y_{t-1}=0$ [16]. As with $w_{i}^{A}$, the (time-varying) weights $w_{it}^{D,1}$ yield a sample from a pseudopopulation in which the distribution of measured baseline covariates within both $A=1$ and $A=0$reflects that in the HPTN 073 population (among individuals who have not yet tested positive for HIV at time $t$). By reweighting the data in the HPTN 061 sample to have the same distribution of baseline covariates as the HPTN 073 population, we (i) address confounding of the $D_{t}$-$Y_{t}$ association due to the causal association between $\boldsymbol{X}_{0}$ and $A$ and (ii) allow estimation of the effect of $D$ in the HPTN 073 population, the target of inference (see DAG in Figure 1) [13].

The inverse probability weights $w_{it}^{D,2}$ yield a sample from a pseudopopulation in which the distribution of the measured covariate history is independent of $D_{t}$, and the distribution of the covariates in each level of $D_{t}$ is the same as that in the entire HPTN 073 population. The weights $w_{i2}^{D,2}$for the HPTN 073 sample address confounding of the $D_{2}$-$Y_{2}$ association by $\boldsymbol{X}_{1}$ and do not need to include $\boldsymbol{X}_{0}$ because $Y_{2}(d_{2})$ and $D_{2}$are independent conditional on $\boldsymbol{X}_{1}$ and $Y_{1}=0$. Such an independence is implied by the DAG in Figure 1 [22] and explicitly described in assumption B3 below.

The weights $w_{it}^{D}$create a sample from a pseudopopulation in which, at each time point $t=1, 2$ and among individuals at risk for HIV, both (i) the distribution of the covariates $\boldsymbol{X}_{0}$ within $A=0$ and (ii) the distribution of the covariates $\boldsymbol{X}_{t-1}$within $A=1$ for each level of $D_{t}$are equivalent to the relevant covariate distributions in the HPTN 073 population, the target of inference [13, 23, 24]. Thus, the weights address all sources of measured confounding of the association between $D$ and HIV risk per the DAG in Figure 1.

We used CBPS to estimate $\Pr\left( A_{i}=1 | \boldsymbol{X}_{i0}, Y_{i,t-1}=0 \right), t=1, 2$ and $\Pr\left( D_{it}=d_{it} | A_{i}=1, \boldsymbol{X}_{t-1},Y_{i,t-1}=0 \right), d_{t}=0, 1, 2, t=1, 2$, using separate models for each time point (i.e., four models were fit). While the levels of $D$ have a natural ordering, we modeled $D$ as a nominal variable when implementing CBPS, which allows for greater flexibility with little loss of information given that $D$ has only three levels.

The model presented in (4),

$$\log\left[ \Pr\left( Y_{t}=1 | D_{t}, Y_{t-1}=0 \right) \right]=\beta_{0t}+\beta_{1}D_{t},$$

is a simplified version of a marginal structural model, which parameterizes the marginal causal effect of a time-varying exposure on a particular outcome as a marginal contrast in potential outcomes [25, 26]. Marginal structural models are often used alongside IPW to account for time-varying covariates, addressing confounding while avoiding conditioning [16, 26, 27].

The weights $w_{it}^{D}$ allowed us to estimate the causal effect of $D$ in the HPTN 073 population, using data from both the HPTN 061 sample and the HPTN 073 sample, provided several key assumptions hold [9, 23]:

(a) Conditional exchangeability for $A$: ${\{Y}_{t}\left( a=0 \right)\perp A|\boldsymbol{X}_{0}, Y_{t-1}=0\}$ for $t=1, 2$ (assumption B1)
(b) Positivity for $A$: $\Pr\left( A=0 | \boldsymbol{X}_{0}=\boldsymbol{x}_{0}\boldsymbol{,}Y_{t-1}=0 \right)>0 \forall\boldsymbol{x}_{0}$for $t=1, 2$ (assumption B2)
(c) Conditional exchangeability for $D$: $\{Y_{t}\left( d_{t} \right)\perp{1(D}_{t}=d_{t})|\boldsymbol{X}_{t-1},A=1,Y_{t-1}=0\}$ for $d_{t}\in\left\{ 0, 1, 2 \right\}, t=1, 2$ (assumption B3)
(d) Positivity for $D$: $0<\Pr\left( D_{t}=d_{t} | A=1, \boldsymbol{X}_{t-1}=\boldsymbol{x}_{t-1},Y_{t-1}=0 \right)<1 \forall\boldsymbol{x}_{t-1},$ for $d_{t}\in\{0, 1, 2\}$, $t=1, 2$ (assumption B4)

The assumption (B1) serves the same role as the assumption (A1) in the $A$ analysis; that is, it allows us to use the observed outcomes for $A=0$ (i.e., $Y_{t}$ in the HPTN 061 population) in place of the potential outcomes $Y_{t}(a=0)$ for $A=1$ (after weighting) [13]. As $A$ has no direct effect on $Y_{t}$ (see Figure 1) and $A=0\Rightarrow D_{t}=0, t=1, 2$, we have $Y_{t}\left( a=0 \right)=Y_{t}(d_{t}=0)$, so we can write (B1) as ${\{Y}_{t}\left( d_{t}=0 \right)\perp A|\boldsymbol{X}_{0}, Y_{t-1}=0\}$, allowing us to use the observed outcomes in the HPTN 061 sample as potential outcomes $Y_{t}(d_{t}=0)$ for $A=1$, that is, in the HPTN 073 population. This also implies that the denominator of $\frac{E[Y_{t}(d_{t})|A=1,Y_{t-1}=0]}{E[Y_{t}\left( d_{t}=0 \right)|A=1,Y_{t-1}=0]}$ is the risk of HIV in the absence of PrEP initiation, regardless of whether there was access to PrEP. Likewise, the assumption of positivity for $A$ (B2) makes $Y_{t}(d_{t}=0)$ realizable for all participants which allows estimation of $E\{Y_{t}(d_{t}=0)|A=1\}$ [13], mirroring assumption (A2).

We further assume exchangeability for $D$ (B3), which allows us to use the observed outcomes for one exposure group (i.e., level of $D$) in place of the potential outcomes for another exposure group within the HPTN 073 population, thus making the causal effect identifiable [13, 28]. This is an assumption of no unmeasured confounding of the association between PrEP initiation/adherence and HIV risk [14, 15]. Lastly, we make an assumption of positivity for $D$ (B4); in particular, we must assume that all individuals in the HPTN 073 population had the potential to have any value of $D$. This assumption makes each potential outcome $Y_{t}(d_{t})$, $d_{t}=0, 1, 2, t=1, 2$ realizable for all participants in the HPTN 073 population and so allows us to estimate the causal effect (3) [11, 13]. As with (A2), if $\Pr\left( D_{t}=d_{t} \right)\approx0$ for the observed value $d_{t}$ for a given observation, then that observation will have an extremely large weight, and could unduly influence the results and decrease precision. While (B1-B4) were the key assumptions, as above we must also assume consistency ($D_{t}=d_{t}\Rightarrow Y_{t}=Y_{t}\left( d_{t} \right)$ for a particular subject) and SUTVA.

In addition to the assumptions enumerated above for the analyses of $A$ and $D$, we must assume (i) no model misspecification in the models $\log\left[ \Pr\left( Y_{2}=1 | A \right) \right]=\beta_{0}+\beta_{1}A$ and $\log\left[ \Pr\left( Y_{t}=1 | D_{t}, Y_{t-1}=0 \right) \right]=\beta_{0t}+\beta_{1}D_{t},$ (ii) no model misspecification in the exposure models for $\Pr\left( A=0 | \boldsymbol{X}_{0}, Y_{t-1}=0 \right)$ and $\Pr\left( D_{t}=d_{t} | A=1, \boldsymbol{X}_{t-1},Y_{t-1}=0 \right), t=1, 2$, and (ii) no measurement error in the exposures, covariates, and outcome [12, 16, 29].

*Missing Data & Confidence Interval Estimation*

Loss to follow-up led to missing data on HIV status for a small fraction of HPTN 073 participants and a larger proportion of HPTN 061 participants. PrEP adherence, which was measured at the six- and twelve-month visits, was available for most individuals who had initiated PrEP. For those for whom it was not available, we attempted to use information on PrEP holds and discontinuations, which were recorded in HPTN 073, to impute the missing values. For all remaining missing data, multiple imputation with chained random forests and predictive mean matching [30] with 10 imputations was implemented using the missRanger package in R [31] in order to include all enrolled participants in the analysis. This approach to missingness assumes the data are missing at random (that is, missingness is random, conditional on observed data) and that the imputation model is correctly specified [32, 33]. The imputation models included the exposures, outcome (HIV status), and covariates used in the main analysis, as well as other variables associated with the exposures and/or outcome.

In particular, the variables included in the multiple imputation procedure are listed in Table S1. Note that the outcome (HIV status) was included in the imputation model, which has been shown to result in improved performance [32, 33]. While some have recommended a “multiple imputation, then deletion” (MID) approach for participants with missing outcomes, wherein the outcome is used in the imputation procedure but imputed outcomes are deleted prior to analysis [60], others have shown that this can produce bias when auxiliary variables used in the imputation are associated with missingness in the outcome [61]. Thus, we did not exclude individuals with imputed outcomes. The data were imputed in “wide” format, the recommended approach for longitudinal data [62].

To combine multiple imputation with inverse probability weights, e.g., for a marginal structural model, the recommended approach is to estimate both the weights and the causal effect in each imputed dataset and then combine these individual effect estimates across datasets to obtain an estimate of the causal effect [32]. We pursued this strategy in our analysis.

**Table S1:** Variables included in multiple imputation procedure.

| **Name** | **Timepoint(s),** $t$ |
| --- | --- |
| Study (HPTN 061 vs. HPTN 073) | 0 |
| HIV status | 1, 2 |
| PrEP initiation | 1, 2 |
| PrEP adherence | 1, 2 |
| STI diagnosis | 0, 1, 2 |
| Number of male partners | 0, 1, 2 |
| URAI | 0, 1, 2 |
| Sexual orientation (gay, bisexual, other) | 0 |
| Region | 0 |
| Age | 0 |
| Primary male partner | 0, 1, 2 |
| Gave money/goods for sex | 0, 1, 2 |
| Received money/goods for sex | 0, 1, 2 |
| HIV positive or unknown male partners | 0, 1, 2 |
| UIAI | 0, 1, 2 |
| URAI with HIV positive or unknown male partners | 0, 1, 2 |
| UIAI with HIV positive or unknown male partners | 0, 1, 2 |
| Use of stimulants (cocaine, methamphetamines) | 0, 1, 2 |
| Use of poppers (inhaled nitrates) | 0, 1, 2 |
| Use of marijuana | 0, 1, 2 |
| Use of heroin | 0, 1, 2 |
| Use of Vicodin | 0, 1, 2 |
| Unprotected sex with alcohol use | 0, 1, 2 |
| Protected sex with alcohol use | 0, 1, 2 |
| Unprotected sex with stimulant use | 0, 1, 2 |
| Protected sex with stimulant use | 0, 1, 2 |
| Unprotected sex with popper use | 0, 1, 2 |
| Protected sex with popper use | 0, 1, 2 |
| Unprotected sex with marijuana use | 0, 1, 2 |
| Protected sex with marijuana use | 0, 1, 2 |
| Trust in healthcare providers | 0, 1, 2 |
| Depression (based on CES-D) [63] | 0, 1, 2 |
| HIV conspiracy beliefs [64, 65] | 0, 1, 2 |
| Internalized homophobia [66] | 0, 1, 2 |
| Social support [67] | 0, 1, 2 |
| Internalized HIV stigma [68] | 0, 1, 2 |
| Alcohol use (based on AUDIT) [69] | 0, 1, 2 |
| Education level | 0 |
| Employment status | 0 |
| Latino/Hispanic | 0 |
| Health insurance coverage | 0 |
| Housing status | 0 |
| Marital status | 0 |
| Income category | 0 |
| Did not have enough money for expenses | 0 |
| Spent $\geq$ 1 night in jail | 0 |

*Note: STI = sexually transmitted infection; URAI = unprotected (condomless) receptive anal intercourse; UIAI = unprotected (condomless) insertive anal intercourse; CES-D = Center for Epidemiologic Studies Depression Scale; AUDIT = Alcohol Use Disorders Identification Test.*

To estimate confidence intervals, we generated 1000 bootstrap samples, stratified by study (HPTN 061 vs. HPTN 073). We performed multiple imputation in each sample and averaged over the imputations (on the log scale) to obtain a single estimate in each bootstrap sample [34]. Based on these 1000 estimates, we estimated percentile bootstrap intervals (again on the log scale). Note that four bootstrap samples (0.4%) were removed because $D_{1}=D_{2}=0$ for all HIV cases, which gave extreme and unstable effect estimates in these samples.

*Diagnostics and Sensitivity Analyses*

Check of Balance

Recall that the utility of the weights $w_{i}^{A}$ and $w_{it}^{D}$ rests on the notion that they are functions of balancing scores and thus address confounding by the relevant (measured) covariates [9]; consequently, it was important to ensure that balance on the included covariates was achieved [35]. Balance is typically assessed by calculating standardized differences and ensuring these differences are small, typically less than 10% [35, 36]. For binary exposures, the standardized difference is obtained by dividing the difference in (weighted) means or proportions of each covariate for the two exposure levels is by the standard deviation [36]. For categorical exposures with more than two levels, standardized differences between each pair of exposure levels are calculated for each covariate [37] and typically summarized by taking the maximum over the exposure level pairs for each covariate [38]. Thus, we assessed whether balance between $A=0$and $A=1$ on the covariates $\boldsymbol{X}_{0}$was achieved by the weights $w_{i}^{A}$ and whether balance between $D_{t}=$ 0, 1, and 2 in HPTN 073 on the covariates $\boldsymbol{X}_{t-1}$was achieved by the weights $w_{it}^{D,2}, t=1, 2$.

For the balance checks comparing $A=0$ and $A=1$, the standard deviation used in the denominator of the standardized difference was that in HPTN 073; this is standard practice when the target estimand is the average treatment effect among the treated (ATT), as is the case in our analysis. For the balance checks comparing $D=$ 0, 1, and 2, the standard deviation used was the pooled standard deviation in HPTN 073, as this is the target population in which we sought balance via the weights $w_{it}^{D,2}$; the pooled standard deviation is generally used when the target estimand is the average treatment effect, i.e., the treatment effect in an entire population, not restricted to a particular value of the exposure [35]. Here we present measures of balance (i) in $\boldsymbol{X}_{0}$for $A=0$ vs. $A=1$, (ii) in $\boldsymbol{X}_{0}$ for $D_{1}=$ 0 vs. 1 vs. 2, and (iii) in $\boldsymbol{X}_{1}$ for $D_{2}=$ 0, 1, 2. While it is also important to consider balance in $\boldsymbol{X}_{0}$ for $A=0$ vs. $A=1$ among those with $Y_{1}=0$ as such balance is needed to address confounding in the $D$ analysis, these results were nearly identical to (i) and are not presented here. We used the cobalt package in R to compute balance measures [39].

The balance measures (Figure S1) correspond to the maximum standardized differences (i.e., the maximum imbalance) across all imputations. We considered the maximum across imputations as it is necessary to ensure balance in each imputed dataset [32]. The measures indicate the presence of covariate imbalance for all three exposures ($A$, $D_{1}$, and $D_{2}$) prior to weighting, and minimal covariate imbalance after weighting.

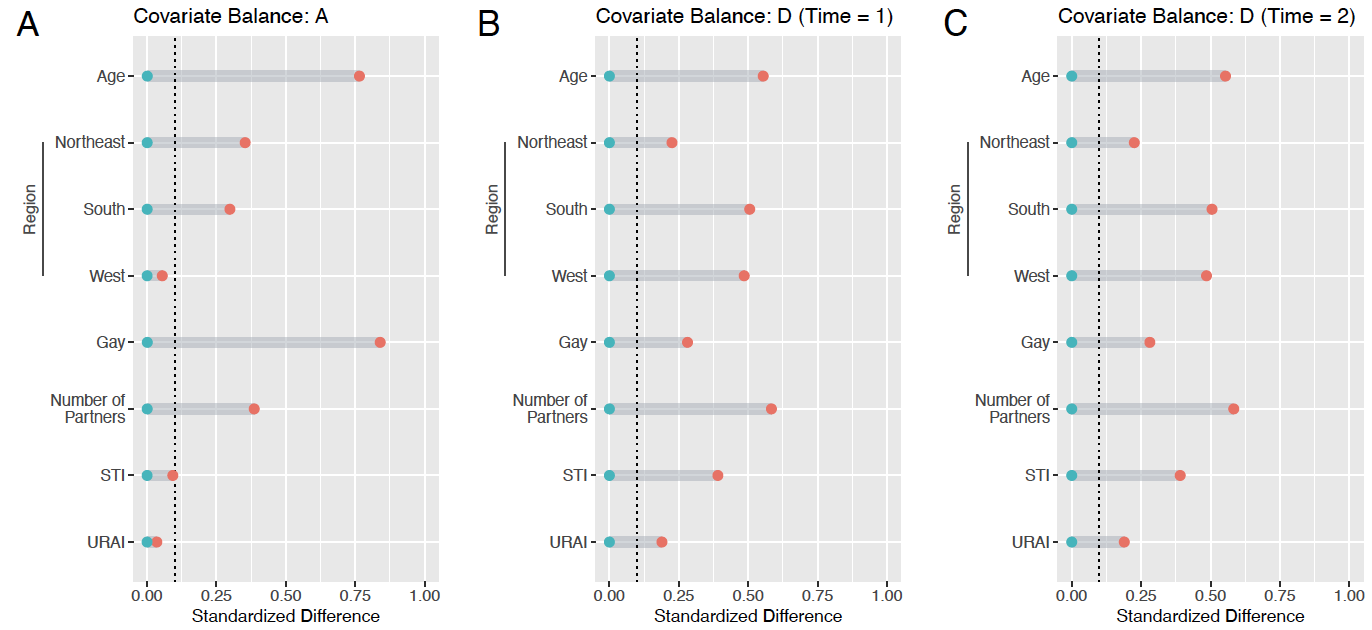

**Figure S1:** Measures of covariate balance before (orange) and after (blue) weighting. These plots summarize the standardized difference across exposure levels for the covariates included in the weight models for three exposures: $A$ (panel A), $D_{1}$ (panel B), and $D_{2}$(panel C). The vertical line at 0.1 highlights the level of imbalance that is generally considered acceptable (10%).

*Note:* STI = sexually transmitted infection; URAI = unprotected (condomless) receptive anal intercourse.

We also considered the maximum degree of covariate imbalance across imputations for each bootstrap sample, as balance is needed in each bootstrap sample in order to obtain valid effect estimates in each sample and thus valid confidence intervals. For the $A$ analysis, covariate balance was achieved through weighting in all bootstrap samples. For $D_{1}$, in 18 bootstrap samples, covariate balance was not achieved after weighting in at least one imputation. For $D_{2}$, covariate imbalance was observed after weighting in 106 bootstrap samples. Consequently, effect estimates from these samples may be biased due to residual confounding by the imbalanced covariate(s), which may in turn affect the validity of the bootstrap confidence intervals. We re-estimated the confidence intervals after removing these 124 samples and observed little change (95% CI for $D=1$vs. $D=0$: 0.130, 0.893; 95% CI for $D=2$vs. $D=0$: 0.017, 0.798).

Check of Positivity

In our analysis, the maximum value of $w_{i}^{A}$ across all bootstrap samples and all imputations was 8.05, while the maximum value of $w_{it}^{D}$ was 159.75; however, consideration of only the weight distribution can mask positivity violations [40]. A better way to identify violations of positivity assumptions is to consider the estimated exposure probabilities, $\hat{\Pr} \left( A=0 | \boldsymbol{X}_{0}, Y_{t-1}=0 \right)$ and $\hat{\Pr} \left( D_{t}=d_{t} | {A=1,\boldsymbol{X}}_{t-1},Y_{t-1}=0 \right)$, and the degree to which these are “extreme.” We note that the estimates of $Pr\left( A=0 | \boldsymbol{X}_{0},Y_{1}=0 \right)$ were nearly identical to the estimates of $\hat{Pr}\left( A=0 | \boldsymbol{X}_{0} \right)$, so only the latter are discussed here. We defined “extreme estimated probabilities” as $\hat{Pr}\left( A=0 | \boldsymbol{X}_{0} \right)<0.001$ and $\hat{Pr}\left( D_{t}=d | {A=1,\boldsymbol{X}}_{t-1},Y_{t-1}=0 \right)<0.001$ or $>0.999, t=1, 2, d=0, 1, 2$. In the original sample, there were very few extreme estimates of exposure probabilities across the 10 imputations (Table S2), with the exception of $\hat{Pr}\left( D_{2}=0 | {A=1,\boldsymbol{X}}_{1},Y_{1}=0 \right)$, for which we observed extreme values in up to 3.2% of individuals in each imputed dataset. These extreme values tended to be close to zero (Figure S2).

Considering the bootstrap samples, we observed that (near-)positivity violations were most common for $\hat{Pr}\left( D_{2}=0 | {A=1,\boldsymbol{X}}_{1},Y_{1}=0 \right)$, with bootstrap samples having up to 49.8% of participants in each imputed dataset with extreme estimated probabilities (Table S2). There was a fairly high proportion of extreme values of $\hat{Pr}\left( D_{2}=1 | {A=1,\boldsymbol{X}}_{1},Y_{1}=0 \right)$ and $\hat{Pr}\left( D_{2}=2 | {A=1,\boldsymbol{X}}_{1},Y_{1}=0 \right)$, as well as $\hat{Pr}\left( D_{1}=0 | {A=1,\boldsymbol{X}}_{0},Y_{1}=0 \right)$ and $\hat{Pr}\left( D_{1}=1 | {A=1,\boldsymbol{X}}_{0},Y_{1}=0 \right)$. After excluding the most extreme 5% of bootstrap samples, the issues with extreme estimates generally only persisted for $\hat{Pr}\left( D_{2}=0 | {A=1,\boldsymbol{X}}_{1},Y_{1}=0 \right)$ and $\hat{Pr}\left( D_{2}=2 | {A=1,\boldsymbol{X}}_{1},Y_{1}=0 \right)$ (Table S2). In general, very small estimated probabilities were observed more often than very large estimated probabilities (Figure S2).

The extreme values observed for $\hat{Pr}\left( D_{2}=0 | {A=1,\boldsymbol{X}}_{1},Y_{1}=0 \right)$ and $\hat{Pr}\left( D_{2}=2 | {A=1,\boldsymbol{X}}_{1},Y_{1}=0 \right)$ were driven by men who were at quite high risk of HIV (and so were more likely to initiate/adhere to PrEP) and at very low risk of HIV (and thus perhaps less motivated to initiate/adhere to PrEP), respectively. In particular, the men in the former group tended to have 4 or more male partners, have an STI, and engage in URAI. The latter group, on the other hand, typically had fewer partners, did not engage in URAI, and did not have STIs. Taken together, these results suggest that for men in the HPTN 073 population with very high levels of risk, the counterfactual $Y_{2}(d_{2}=0)$ may not be realizable, and for men with very low levels of risk, the counterfactuals $Y_{2}(d_{2}=1)$ and $Y_{2}(d_{2}=2)$ might not be realizable. Consequently, our estimates of the effect of PrEP initiation and adherence in these very high and very low risk men (including the estimates from the bootstrap samples, which were used to estimate confidence intervals) may be based on extrapolation [41, 42].

Importantly, others have noted that near-violations of positivity generally must be extreme to have a practical impact on the causal effect estimate [43]. Furthermore, the possible impact of near-violations of positivity in our analysis mainly affects the bootstrap confidence intervals; given that extreme weights generally result in high variability, our confidence intervals are likely to be conservative.

**Table S2:** Proportion of extreme estimates for each exposure probability, across imputations in the original sample and across bootstrap samples and imputations.

| **Probability** | **Original sample** | **Bootstrap** | |
| --- | --- | --- | --- |
|  | **Max.**$\dagger$ | **Max.**$\ddagger$ | **95^th^ percentile**$\ddagger$ |
| $\Pr\left( A=0 \vert\boldsymbol{X}_{0} \right)$ | 0% | 0% | 0% |
| $\Pr\left( D_{1}=0 \vert{A=1, \boldsymbol{X}}_{0} \right)$ | 0% | 18.1% | 1.3% |
| $\Pr\left( D_{1}=1 \vert{A=1, \boldsymbol{X}}_{0} \right)$ | 0% | 9.3% | 0% |
| $\Pr\left( D_{1}=2 \vert{A=1, \boldsymbol{X}}_{0} \right)$ | 0% | 0.9% | 0% |
| $\Pr\left( D_{2}=0 \vert{A=1, \boldsymbol{X}}_{1},Y_{1}=0 \right)$ | 3.2% | 49.8% | 37.7% |
| $\Pr\left( D_{2}=1 \vert{A=1, \boldsymbol{X}}_{1},Y_{1}=0 \right)$ | 0% | 9.2% | 0.5% |
| $\Pr\left( D_{2}=2 \vert{A=1, \boldsymbol{X}}_{1},Y_{1}=0 \right)$ | 0% | 14.7% | 6.4% |

$\dagger$Maximum proportion of extreme estimates across imputations. Extreme observations for $\Pr\left( A=0 | \boldsymbol{X}_{0} \right)$are those < 0.001; extreme observations for the remainder of the probabilities are those < 0.001 or > 0.999. The proportion of extreme observations is computed for each imputation, and the maximum across imputations is computed. $\ddagger$Maximum and 95^th^ percentile proportion of extreme observations across bootstrap samples and imputations.

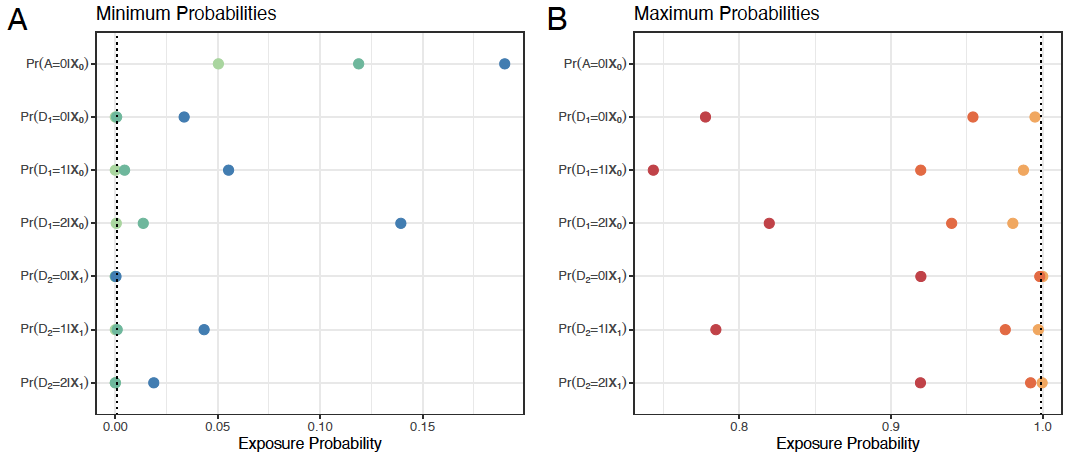

**Figure S2:** Summary of extreme values of estimated exposure probabilities across imputations and across bootstrap samples and imputations. Minimum values are show in panel A and maximum values are shown in panel B. In panel A, the minimum across 10 imputations in the original sample is shown in dark blue, the 5^th^ percentile across bootstrap samples of the minimum probability among imputations in each bootstrap sample is shown in dark green, and the minimum across all bootstrap samples and all imputations is shown in light green. In panel B, the maximum across 10 imputations in the original sample is shown in dark red, the 95^th^ percentile across bootstrap samples of the maximum probability among imputations in each bootstrap sample is shown in dark orange, and the maximum across all bootstrap samples and all imputations is shown in light orange. Note that $P(A=0|\boldsymbol{X}_{0})$ are not shown in panel B because no assumptions were made regarding the maximum value of $P(A=0|\boldsymbol{X}_{0})$. The vertical lines indicate extreme probabilities (defined as falling below 0.001 or above 0.999).

Unmeasured Confounding

Unmeasured or uncontrolled confounding (referred to as “unmeasured confounding” hereafter) arises from incomplete adjustment of one or more confounders, either because they are unmeasured, they are measured with error, or there is residual confounding due to improper adjustment. We evaluated the potential impact of unmeasured confounding on our casual effect estimates using methods similar to those of Robins [44], Brumback et al. [45], Chiba [46], and Klungsøyr et al. [47] (see derivation of our implementation below), which can be used to estimate the true causal effect of an exposure given a certain degree of unmeasured confounding. The magnitude and direction of unmeasured confounding is quantified by the bias factor, here defined for $A:$

$$\alpha=\frac{\Pr\left( Y_{2}\left( a \right)=1 | A=1, \boldsymbol{X}_{0} \right)}{\Pr\left( Y_{2}\left( a \right)=1 | A=0, \boldsymbol{X}_{0} \right)},$$

which was assumed to be the same for $a\in\left( 0, 1 \right)$and to not depend on $\boldsymbol{X}_{0}$[46]. Chiba [46] proposed considering

$$\frac{1}{RR_{c}}\leq\alpha\leq RR_{c},$$

where $RR_{c}>1$ is the RR of $Y_{2}$ for a strong risk factor; we fixed $RR_{c}=2$. We observed that for $\alpha=$0.71, the estimated RR of HIV for $A=1$vs. $A=0$would be 0.77 (95% CI: 0.32, 1.78) (Figure S3). Conversely, for $\alpha=$1.4, the estimated RR of HIV for PrEP access would be 0.35 (0.14, 0.82).

Likewise for the analysis of PrEP initiation/adherence ($D$), the bias factor is defined as

$$\alpha_{t}\left( d, d^{'} \right)=\frac{Pr(Y_{t}\left( d \right)=1|D_{t}=d^{'},\boldsymbol{X}_{t-1})}{Pr(Y_{t}\left( d \right)=1|D_{t}=d, \boldsymbol{X}_{t-1})},$$

for $d, d^{'}\in\{0, 1, 2\}$ and $t=1, 2$. We assumed $\alpha_{t}\left( d, d^{'} \right)$does not depend on $\boldsymbol{X}_{t-1}$and $\alpha_{1}\left( d,d^{'} \right)=\alpha_{2}\left( d,d^{'} \right)\equiv\alpha\left( d,d^{'} \right) \forall d, d'$. In addition, we assumed $\alpha\left( 0,1 \right)=\alpha\left( 1,2 \right), \alpha\left( 0,2 \right)=\alpha\left( 0,1 \right)^{2},$and $\alpha\left( d,d^{'} \right)=1/\alpha(d^{'},d)$. We considered

$$\frac{1}{RR_{c}}\leq\alpha\left( 0,1 \right)\leq RR_{c},$$

and $RR_{c}=2$; $RR_{c}$ can be conceptualized as above. We found that for $\alpha\left( 0,1 \right)=$0.71, the estimated RR of HIV for $D=1$ vs. $0$ was 0.66 (95% CI: 0.17, 1.15) and the estimated RR of HIV for $D=2$ vs. 0 was 0.43 (0.03, 1.33). For $\alpha\left( 0,1 \right)=$1.4, these estimated RRs were 0.34 (0.09, 0.70) and 0.12 (0.01, 0.49), respectively.

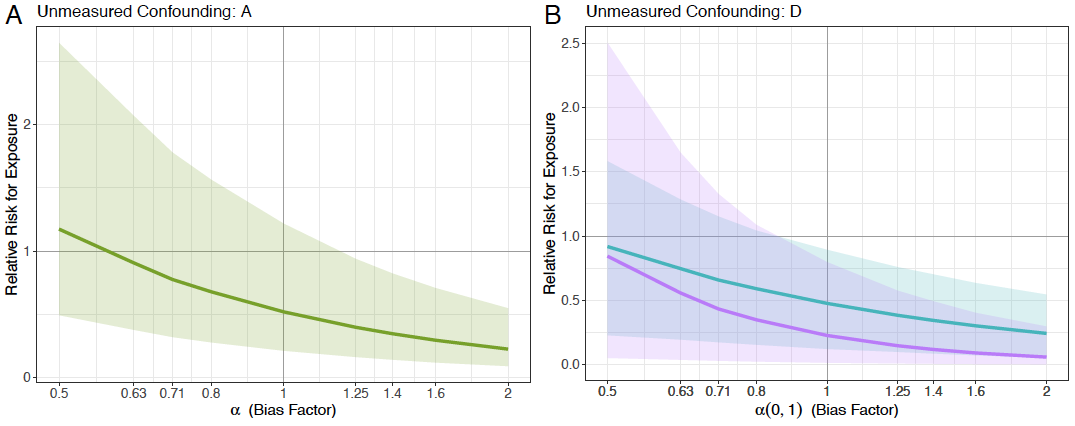

**Figure S3:** Results of the sensitivity analysis to assess the impact of unmeasured confounding. Estimates of the true causal effect of PrEP access (panel A) and PrEP initiation/adherence (panel B) and corresponding 95% confidence intervals were obtained using the bias factor (x-axis) to quantify the effect of unmeasured confounding. In panel B, blue indicates D = 1 vs. 0, while purple corresponds to D = 2 vs. 0.

*Alternative Approaches*

Our approach, based on marginal structural models with probability weighting, is quite common, though alternatives exist. Perhaps the most obvious is a standard regression model, i.e., a generalized linear model adjusting for the suspected confounding factors. This would have provided an estimate of the conditional effects of PrEP access, initiation, and adherence [27], which may differ from the marginal effect in the presence of effect modification [48, 49]. Furthermore, such an analysis would be computationally difficult given our very rare outcome and the need to adjust for a non-trivial number of presumed confounders. Conversely, estimating the weights in our analysis was straightforward as the exposures were quite common. Additionally, while our analysis relies on strong assumptions, similar assumptions (including no measurement error, no unmeasured confounding, no model misspecification, and no interference) are needed for the estimates from a standard regression model to have a causal interpretation [12, 16, 29]. Our analytic approach also allowed us to be very deliberate about our target of inference. A regression model, on the other hand, would provide estimates for a population that is an ambiguous weighted average of the HPTN 061 and HPTN 073 populations [50].

We have alluded to the fact that a randomized, placebo-controlled trial may be useful in estimating the causal effect of PrEP in Black MSM, but such a trial would be unethical in our setting. Furthermore, it is not feasible to randomize individuals to levels of PrEP adherence, or even to PrEP adherence or non-adherence: at most, participants can be provided with pills and instructions and their adherence monitored, but they cannot be forced to take medication (though there are biobehavioral interventions that can improve PrEP adherence, e.g., [51]). Marginal structural models, which can be thought of as seeking to approximate an unblinded randomized trial [52, 53], are thus particularly useful here. Importantly, however, as discussed in the main paper, our approach can provide balance between exposure levels only on measured variables [17]. This is in contrast with randomized trials, which provide balance on all variables (in expectation). On the other hand, while randomization yields balance in expectation on measured and unmeasured variables, allowing estimation of the causal effect, the presence of non-representative sampling [7, 54, 55], non-compliance [21, 53], chance imbalances [7, 55], and/or loss to follow-up [12] in randomized trials can threaten both internal and external validity. In addition, given the challenges related to recruiting stigmatized populations, like Black MSM, for research, it may be difficult to conduct a randomized trial in this population [8] and the careful use of existing data and multiple data sources (e.g., external controls) can yield valuable insights.

It is common to assume a parsimonious dose-response relationship [26] for a non-binary exposure, as we have done in the analysis of $D$. While it would have been preferable to fit a more flexible model, doing so was not possible as there were no cases of HIV among individuals with $D=2$ in the original sample (before imputation). Since $D$ was modeled as an ordinal variable, the estimate of the causal effects of $D=2$ vs. $D=0$ and $D=1$ vs. $D=0$ are constrained, given the presumed log-linear relationship. Given that previous research has found very large reductions in risk of HIV among individuals who were adherent to PrEP [56-58], there is evidence that our analysis, in which the relationship between $D$ and HIV risk is constrained, may have overestimated the effect of PrEP initiation without adherence, and underestimated the effect of PrEP adherence. Indeed, in a sensitivity analysis among those with $D<2$, the magnitude of the estimated relative risk of $D=1$vs. $D=0$, i.e., the effect of PrEP initiation without adherence, was less extreme (RR = 0.60). As a consequence of this attenuated effect and reduced sample size, the 95% CI was shifted and wider (0.11, 2.22).

Another possibility would have been to include datasets from other HPTN studies involving Black MSM in the US, e.g., HPTN 083, which was a randomized study comparing long-acting injectable cabotegravir to daily oral TDF-FTC [59]. The TDF-FTC arm, which was largely comprised of MSM and included 434 Black participants in the US, could have contributed to our analysis. However, we chose not to include data from this study for two reasons. First, HPTN 083 was a randomized study and is thus susceptible to the challenges described above related to external validity. Second, HPTN 083 was conducted between December 2016 and March 2020, making it fairly far removed temporally from HPTN 061.

*Derivations: Weights*

I. $A$ analysis

In HPTN 061,

$$\Pr\left( Y_{2}\left( a=0 \right)=1 | A=1 \right)=\sum_{\boldsymbol{x}_{0}} \Pr\left( Y_{2}\left( a=0 \right)=1 | A=1,\boldsymbol{X}_{0}=\boldsymbol{x}_{0} \right)Pr(\boldsymbol{X}_{0}=\boldsymbol{x}_{0}|A=1)$$

$$=\sum_{\boldsymbol{x}_{0}} \Pr\left( Y_{2}\left( a=0 \right)=1 | A=0,\boldsymbol{X}_{0}=\boldsymbol{x}_{0} \right)\Pr\left( \boldsymbol{X}_{0}=\boldsymbol{x}_{0} | A=1 \right) due to (A1)$$

$$=\sum_{\boldsymbol{x}_{0}} \Pr\left( Y_{2}=1 | A=0,\boldsymbol{X}_{0}=\boldsymbol{x}_{0} \right)\Pr\left( \boldsymbol{X}_{0}=\boldsymbol{x}_{0} | A=1 \right)$$

$$= \sum_{\boldsymbol{x}_{0}} \frac{\Pr\left( Y_{2}=1,A=0,\boldsymbol{X}_{0}=\boldsymbol{x}_{0} \right)}{\Pr\left( A=0,\boldsymbol{X}_{0}=\boldsymbol{x}_{0} \right)}\Pr\left( \boldsymbol{X}_{0}=\boldsymbol{x}_{0} | A=1 \right)$$

$$=\sum_{\boldsymbol{x}_{0}} \frac{Pr(Y_{2}=1,A=0,\boldsymbol{X}_{0}=\boldsymbol{x}_{0})}{Pr(A=0|\boldsymbol{X}_{0}=\boldsymbol{x}_{0})}\frac{Pr(\boldsymbol{X}_{0}=\boldsymbol{x}_{0}|A=1)}{Pr(\boldsymbol{X}_{0}=\boldsymbol{x}_{0})}$$

$$=\frac{1}{Pr(A=1)}\sum_{\boldsymbol{x}_{0}} Pr(Y_{2}=1,A=0,\boldsymbol{X}_{0}=\boldsymbol{x}_{0})\frac{Pr(A=1|\boldsymbol{X}_{0}=\boldsymbol{x}_{0})}{Pr(A=0|\boldsymbol{X}_{0}=\boldsymbol{x}_{0})}$$

$$=\frac{1}{Pr(A=1)}\sum_{\boldsymbol{x}_{0}} \Pr\left( Y_{2}=1,A=0,\boldsymbol{X}_{0}=\boldsymbol{x}_{0} \right)w^{A}(\boldsymbol{x}_{0})$$

as shown in Sato and Matsuyama [70]. Furthermore, as demonstrated in Hernan and Robins [12], we can write

$$\sum_{\boldsymbol{x}_{0}} \Pr\left( Y_{2}=1,A=0,\boldsymbol{X}_{0}=\boldsymbol{x}_{0} \right)w^{A}(\boldsymbol{x}_{0})=E\left[ \Pr\left( Y_{2}=1, A=0 | \boldsymbol{X}_{0} \right)w^{A}\left( \boldsymbol{X}_{0} \right) \right]=E[1\left( A=0 \right)Y_{2}w^{A}\left( \boldsymbol{X}_{0} \right)]$$

using the definition of expectation and

$$\Pr\left( A=1 \right)=E\left[ 1\left( A=0 \right)w^{A}\left( \boldsymbol{X}_{0} \right) \right].$$

Thus, we can write

$$\Pr\left( Y_{2}\left( a=0 \right)=1 | A=1 \right)=\frac{E\left[ 1\left( A=0 \right)Y_{2}w^{A}(\boldsymbol{X}_{0}) \right]}{E[1\left( A=0 \right)w^{A}\left( \boldsymbol{X}_{0} \right)]}$$

where the quantity on the right-hand size is the inverse-probability weighted mean. Consequently, the weights $w^{A}(\boldsymbol{X}_{0})$ can be used to estimate the inverse-probability weighted mean and obtain the correct causal quantity.

Similarly, in HPTN 073,

$$\Pr\left( Y_{2}\left( a=1 \right)=1 | A=1 \right)= \sum_{\boldsymbol{x}_{0}} \Pr\left( Y_{2}\left( a=1 \right)=1 | A=1,\boldsymbol{X}_{0}=\boldsymbol{x}_{0} \right)Pr(\boldsymbol{X}_{0}=\boldsymbol{x}_{0}|A=1)$$

$$=\sum_{\boldsymbol{x}_{0}} \Pr\left( Y_{2}=1 | A=1,\boldsymbol{X}_{0}=\boldsymbol{x}_{0} \right)\Pr\left( \boldsymbol{X}_{0}=\boldsymbol{x}_{0} | A=1 \right)$$

$$= \sum_{\boldsymbol{x}_{0}} \frac{\Pr\left( Y_{2}=1,A=1,\boldsymbol{X}_{0}=\boldsymbol{x}_{0} \right)}{\Pr\left( A=1,\boldsymbol{X}_{0}=\boldsymbol{x}_{0} \right)}\Pr\left( \boldsymbol{X}_{0}=\boldsymbol{x}_{0} | A=1 \right)$$

$$=\sum_{\boldsymbol{x}_{0}} \frac{Pr(Y_{2}=1,A=1,\boldsymbol{X}_{0}=\boldsymbol{x}_{0})}{Pr(A=1|\boldsymbol{X}_{0}=\boldsymbol{x}_{0})}\frac{Pr(\boldsymbol{X}_{0}=\boldsymbol{x}_{0}|A=1)}{Pr(\boldsymbol{X}_{0}=\boldsymbol{x}_{0})}$$

$$=\frac{1}{Pr(A=1)}\sum_{\boldsymbol{x}_{0}} Pr(Y_{2}=1,A=1,\boldsymbol{X}_{0}=\boldsymbol{x}_{0})\cdot1$$

$$=\frac{1}{Pr(A=1)}\sum_{\boldsymbol{x}_{0}} Pr(Y_{2}=1,A=1,\boldsymbol{X}_{0}=\boldsymbol{x}_{0})\cdot w^{A}(\boldsymbol{x}_{0})$$

and

$$\Pr\left( Y_{2}\left( a=1 \right)=1 | A=1 \right)=\frac{1}{Pr(A=1)}\sum_{\boldsymbol{x}_{0}} Pr(Y_{2}=1,A=1,\boldsymbol{X}_{0}=\boldsymbol{x}_{0})=\frac{E\left[ 1\left( A=1 \right)Y_{2}w^{A}(\boldsymbol{X}_{0}) \right]}{E[1\left( A=1 \right)w^{A}\left( \boldsymbol{X}_{0} \right)]}.$$

II. $D$ analysis

Note that these derivations are based on the form of the weights in (4).

In HPTN 061,

$$\Pr\left( Y_{t}\left( d_{t}=0 \right)=1 | A=1,Y_{t-1}=0 \right)=Pr(Y_{t}\left( a=0 \right)=1|A=1,Y_{t-1}=0)=\sum_{\boldsymbol{x}_{0}} \Pr\left( Y_{t}\left( d_{t}= 0 \right)=1 | A=1,\boldsymbol{X}_{\boldsymbol{0}}=\boldsymbol{x}_{0},Y_{t-1}=0 \right)\Pr\left( \boldsymbol{X}_{\boldsymbol{0}}=\boldsymbol{x}_{0} | A=1,Y_{t-1}=0 \right)$$

$$=\sum_{\boldsymbol{x}_{0}} Pr(Y_{t}\left( d_{t}=0 \right)=1\left| A=0,\boldsymbol{X}_{\boldsymbol{0}}=\boldsymbol{x}_{0},Y_{t-1}=0 \right)\Pr\left( \boldsymbol{X}_{\boldsymbol{0}}=\boldsymbol{x}_{0} | A=1,Y_{t-1}=0 \right)from (B1)$$

$$=\sum_{\boldsymbol{x}_{0}} \Pr\left( Y_{t}=1 | A=0,\boldsymbol{X}_{\boldsymbol{0}}=\boldsymbol{x}_{0},Y_{t-1}=0 \right)Pr(\boldsymbol{X}_{\boldsymbol{0}}=\boldsymbol{x}_{0}|A=1,Y_{t-1}=0)$$

$$=\sum_{\boldsymbol{x}_{0}} \frac{Pr(Y_{t}=1,A=0,\boldsymbol{X}_{\boldsymbol{0}}=\boldsymbol{x}_{0},Y_{t-1}=0)}{Pr(A=0, \boldsymbol{X}_{\boldsymbol{0}}=\boldsymbol{x}_{0},Y_{t-1}=0)}Pr(\boldsymbol{X}_{\boldsymbol{0}}=\boldsymbol{x}_{0}|A=1,Y_{t-1}=0)$$

$$=\sum_{\boldsymbol{x}_{0}} \frac{Pr(Y_{t}=1,A=0,\boldsymbol{X}_{\boldsymbol{0}}=\boldsymbol{x}_{0},Y_{t-1}=0)}{Pr(A=0,Y_{t-1}=0|\boldsymbol{X}_{\boldsymbol{0}}=\boldsymbol{x}_{0})}\frac{Pr(\boldsymbol{X}_{\boldsymbol{0}}=\boldsymbol{x}_{0}|A=1,Y_{t-1}=0)}{Pr(\boldsymbol{X}_{\boldsymbol{0}}=\boldsymbol{x}_{0})}$$

$$=\frac{1}{Pr(A=1,Y_{t-1}=0)}\sum_{\boldsymbol{x}_{0}} Pr(Y_{t}=1,A=0,\boldsymbol{X}_{\boldsymbol{0}}=\boldsymbol{x}_{0},Y_{t-1}=0)\frac{Pr(A=1|\boldsymbol{X}_{\boldsymbol{0}}=\boldsymbol{x}_{0},Y_{t-1}=0)}{Pr(A=0|\boldsymbol{X}_{\boldsymbol{0}}=\boldsymbol{x}_{0},Y_{t-1}=0)}$$

$$=\frac{1}{\Pr\left( A=1,Y_{t-1}=0 \right)}\sum_{\boldsymbol{x}_{0}} \Pr\left( Y_{t}=1,A=0,\boldsymbol{X}_{\boldsymbol{0}}=\boldsymbol{x}_{0},Y_{t-1}=0 \right)w_{t}^{D}\left( \boldsymbol{x}_{0} \right),$$

where the first line follows from the fact that $A$ has no direct effect on $Y_{t}$ (see Figure 1) and $A=0\Rightarrow D_{t}=0, t=1, 2,$ so $Y_{t}\left( a=0 \right)=Y_{t}\left( d_{t}=0 \right).$We can further show that

$$\Pr\left( Y_{t}\left( d_{t}=0 \right)=1 | A=1,Y_{t-1}=0 \right)=\frac{1}{\Pr\left( A=1,Y_{t-1}=0 \right)}\sum_{\boldsymbol{x}_{0}} \Pr\left( Y_{t}=1,A=0,\boldsymbol{X}_{\boldsymbol{0}}=\boldsymbol{x}_{0},Y_{t-1}=0 \right)w_{t}^{D}\left( \boldsymbol{x}_{0} \right)= \frac{E\left[ 1\left( A=0,Y_{t-1}=0 \right)Y_{t}w_{t}^{D}\left( \boldsymbol{X}_{\boldsymbol{0}} \right) \right]}{E[1\left( A=0, Y_{t-1}=0 \right)w_{t}^{D}\left( \boldsymbol{X}_{\boldsymbol{0}} \right)]}$$

using similar arguments as above.

In HPTN 073,

$$\Pr\left( Y_{t}\left( d_{t} \right)=1 | A=1,Y_{t-1}=0 \right)=\sum_{\boldsymbol{x}_{t-1}} \Pr\left( Y_{t}\left( d_{t} \right)=1 | A=1,Y_{t-1}=0,\boldsymbol{X}_{\boldsymbol{t-1}}\boldsymbol{=}\boldsymbol{x}_{t-1} \right)Pr(\boldsymbol{X}_{\boldsymbol{t-1}}\boldsymbol{=}\boldsymbol{x}_{t-1}|A=1,Y_{t-1}=0)$$

$$=\sum_{\boldsymbol{x}_{t-1}} \Pr\left( Y_{t}\left( d_{t} \right)=1|D_{t}=d_{t}, A=1, Y_{t-1}=0,\boldsymbol{X}_{\boldsymbol{t-1}}\boldsymbol{=}\boldsymbol{x}_{t-1} \right)Pr(\boldsymbol{X}_{\boldsymbol{t-1}}\boldsymbol{=}\boldsymbol{x}_{t-1}|A=1,Y_{t-1}=0) from (B3)$$

$$=\sum_{\boldsymbol{x}_{t-1}} \Pr\left( Y_{t}=1|D_{t}=d_{t}, A=1, Y_{t-1}=0,\boldsymbol{X}_{\boldsymbol{t-1}}\boldsymbol{=}\boldsymbol{x}_{t-1} \right)Pr(\boldsymbol{X}_{\boldsymbol{t-1}}\boldsymbol{=}\boldsymbol{x}_{t-1}|A=1,Y_{t-1}=0)$$

$$=\sum_{\boldsymbol{x}_{t-1}} \frac{Pr(Y_{t}=1,D_{t}=d_{t}, A=1, Y_{t-1}=0,\boldsymbol{X}_{\boldsymbol{t-1}}\boldsymbol{=}\boldsymbol{x}_{t-1})}{Pr(D_{t}=d_{t}, A=1, Y_{t-1}=0,\boldsymbol{X}_{\boldsymbol{t-1}}\boldsymbol{=}\boldsymbol{x}_{t-1})}Pr(\boldsymbol{X}_{\boldsymbol{t-1}}\boldsymbol{=}\boldsymbol{x}_{t-1}|A=1,Y_{t-1}=0)$$

$$=\sum_{\boldsymbol{x}_{t-1}} \frac{Pr(Y_{t}=1,D_{t}=d_{t}, A=1, Y_{t-1}=0,\boldsymbol{X}_{\boldsymbol{t-1}}\boldsymbol{=}\boldsymbol{x}_{t-1})}{Pr(D_{t}=d_{t}, A=1, Y_{t-1}=0,\boldsymbol{X}_{\boldsymbol{t-1}}\boldsymbol{=}\boldsymbol{x}_{t-1})}\frac{Pr(\boldsymbol{X}_{\boldsymbol{t-1}}\boldsymbol{=}\boldsymbol{x}_{t-1},A=1,Y_{t-1}=0)}{Pr(A=1,Y_{t-1}=0)}$$

$$=\frac{1}{Pr(A=1,Y_{t-1}=0)}\sum_{\boldsymbol{x}_{t-1}} \frac{Pr(Y_{t}=1,D_{t}=d_{t}, A=1, Y_{t-1}=0,\boldsymbol{X}_{\boldsymbol{t-1}}\boldsymbol{=}\boldsymbol{x}_{t-1})}{Pr(D_{t}=d_{t}| A=1, Y_{t-1}=0,\boldsymbol{X}_{\boldsymbol{t-1}}\boldsymbol{=}\boldsymbol{x}_{t-1})}$$

$$=\frac{1}{Pr(A=1,Y_{t-1}=0)}\sum_{\boldsymbol{x}_{t-1}} Pr(Y_{t}=1,D_{t}=d_{t}, A=1, Y_{t-1}=0,\boldsymbol{X}_{\boldsymbol{t-1}}\boldsymbol{=}\boldsymbol{x}_{t-1})w_{t}^{D}(\boldsymbol{x}_{t-1}).$$

Likewise, using similar arguments as above, we can show

$$\Pr\left( Y_{t}\left( d_{t} \right)=1 | A=1,Y_{t-1}=0 \right)=\frac{1}{Pr(A=1,Y_{t-1}=0)}\sum_{\boldsymbol{x}_{t-1}} \Pr\left( Y_{t}=1,D_{t}=d_{t}, A=1, Y_{t-1}=0,\boldsymbol{X}_{\boldsymbol{t-1}}\boldsymbol{=}\boldsymbol{x}_{t-1} \right)w_{t}^{D}\left( \boldsymbol{x}_{t-1} \right)= \frac{E\left[ 1\left( D_{t}=d_{t},Y_{t-1}=0,A=1 \right)Y_{t}w_{t}^{D}\left( \boldsymbol{X}_{\boldsymbol{t-1}} \right) \right]}{E\left[ 1\left( D_{t}=d_{t}, Y_{t-1}=0,A=1 \right)w_{t}^{D}\left( \boldsymbol{X}_{\boldsymbol{t-1}} \right) \right]}.$$

*Derivations: Sensitivity Analysis for Unmeasured Confounding*

I. $A$ analysis

Using the nomenclature and notation of Chiba [46], we define the bias factors as

$$\alpha=\frac{\Pr\left( Y_{2}\left( a=1 \right)=1 | A=1, \boldsymbol{X}_{0} \right)}{\Pr\left( Y_{2}\left( a=1 \right)=1 | A=0, \boldsymbol{X}_{0} \right)}$$

and

$$\beta= \frac{\Pr\left( Y_{2}\left( a=0 \right)=1 | A=1, \boldsymbol{X}_{0} \right)}{\Pr\left( Y_{2}\left( a=0 \right)=1 | A=0, \boldsymbol{X}_{0} \right)},$$

which we assume do not depend on $\boldsymbol{X}_{0}$. If there are no unmeasured confounders and thus (A1) holds, then $\alpha=\beta=1$. If there is an unmeasured confounder, then $\alpha$ is the relative risk for $A$ of $Y_{2}(a=1)$, conditional on $\boldsymbol{X}_{0}$, and $\beta$ is the relative risk for $A$ of $Y_{2}\left( a=0 \right)$, conditional on $\boldsymbol{X}_{0}$[45]. Thus, the bias factors capture the magnitude and direction of unmeasured confounding. If the unmeasured confounder is binary and collinear with $A$ (i.e., an extreme form of confounding), then $\alpha$can be conceptualized as the relative risk of the unmeasured confounder for $Y_{2}(a=1)$ and $\beta$ is the relative risk of the unmeasured confounder for $Y_{2}(a=0)$ (conditional on $\boldsymbol{X}_{0}$).

Based on these bias factors, we can define the bias-corrected versions of $Y_{2}$:

$$Y_{2}^{\alpha}=\left\{ \Pr\left( A=1 | \boldsymbol{X}_{0} \right)+\frac{Pr(A=0|\boldsymbol{X}_{0})}{\alpha} \right\}Y_{2}$$

and

$$Y_{2}^{\beta}=\left\{ \beta\Pr\left( A=1 | \boldsymbol{X}_{0} \right)+Pr(A=0|\boldsymbol{X}_{0}) \right\}Y_{2}.$$

Importantly, these bias factors are consistent: if $\alpha=1$, then $Y_{2}^{\alpha}=Y_{2}$ and if $\beta=1$, then $Y_{2}^{\beta}=Y_{2}$. Furthermore,

$$E\left[ Y_{2}^{\alpha} | A=1, \boldsymbol{X}_{0} \right]=E[\left\{ \Pr\left( A=1 | \boldsymbol{X}_{0} \right)+\Pr\left( A=0 | \boldsymbol{X}_{0} \right)/\alpha\}Y_{2}] | A=1, \boldsymbol{X}_{0} \right]= E\left[ \left. \Pr\left( A=1 | \boldsymbol{X}_{0} \right)Y_{2}+\frac{\Pr\left( A=0 | \boldsymbol{X}_{0} \right)\Pr\left( Y_{2}\left( a=1 \right)=1 | A=0, \boldsymbol{X}_{0} \right)}{\Pr\left( Y_{2}\left( a=1 \right)=1 | A=1,\boldsymbol{X}_{0} \right)}Y_{2} \right|A=1, \boldsymbol{X}_{0} \right]= E\left[ \left. \frac{\Pr\left( Y_{2}\left( a=1 \right)=1 | \boldsymbol{X}_{0} \right)}{\Pr\left( Y_{2}\left( a=1 \right)=1 | A=1,\boldsymbol{X}_{0} \right)}Y_{2} \right|A=1,\boldsymbol{X}_{0} \right]= \frac{\Pr\left( Y_{2}\left( a=1 \right)=1 | \boldsymbol{X}_{0} \right)\Pr\left( Y_{2}=1 | A=1,\boldsymbol{X}_{0} \right)}{\Pr\left( Y_{2}\left( a=1 \right)=1 | A=1,\boldsymbol{X}_{0} \right)} = \frac{\Pr\left( Y_{2}\left( a=1 \right)=1 | \boldsymbol{X}_{0} \right)\Pr\left( Y_{2}\left( a=1 \right)=1 | A=1,\boldsymbol{X}_{0} \right)}{\Pr\left( Y_{2}\left( a=1 \right)=1 | A=1,\boldsymbol{X}_{0} \right)} by consistency =\Pr\left( Y_{2}\left( a=1 \right)=1 | \boldsymbol{X}_{0} \right).$$

Thus, the use of $Y_{2}^{\alpha}$ removes the bias from unmeasured confounding given $\alpha$, providing an unbiased estimate of the causal relative risk. Likewise, $E\left[ Y_{2}^{\beta}|A=0, \boldsymbol{X}_{0} \right]=\Pr(Y_{2}\left( a=0 \right)=1|\boldsymbol{X}_{0}),$ meaning that $Y_{2}^{\beta}$ removes the bias from unmeasured confounding given $\beta$.

In our sensitivity analysis, we assumed that $\alpha=\beta$, as in Chiba [46]. Chiba [46] considered

$$\frac{1}{RR_{c}}\leq\alpha=\beta\leq RR_{c},$$

where $RR_{c}>1$ is the relative risk of $Y_{2}$ for a strong risk factor. We evaluated $\alpha=\beta=RR_{c}$ for $RR_{c}\in\{0.5, 0.63, 0.71, 0.8, 1, 1.25, 1.4, 1.6, 2\}$.

II. $D$ analysis

Note that these derivations are based on the form of the weights in (4).

Relying on the notation of Klungsøyr et al. [47], and extending their methods to repeated measures and the relative risk (as opposed to the hazard ratio), we can define the bias factors due to unmeasured confounding as:

$$\gamma\left( d,t \right)=\frac{\sum_{j=0}^{2} \left[ \Pr\left( D_{t}=j \right|\boldsymbol{X}_{t-1})Pr(Y_{t}\left( d \right)=1|D_{t}=j,\boldsymbol{X}_{t-1}) \right]}{\Pr\left( Y_{t}\left( d \right)=1 | D_{t}=d, \boldsymbol{X}_{t-1} \right)}=\sum_{j=0}^{2} [\Pr\left( D_{t}=j | \boldsymbol{X}_{t-1} \right)\alpha_{t}\left( d, j \right)],$$

where $d=0, 1, 2, t=1, 2$. We assume these bias factors do depend on $\boldsymbol{X}_{t-1}$. If there is no unmeasured confounding, then $\gamma(d,t)=1 \forall d, t$. We can use these bias factors to define bias-corrected versions of $Y_{t}$:

$$Y_{t}^{\gamma\left( d,t \right)}=\gamma\left( d,t \right)Y_{t}.$$

We can also demonstrate that the use of $Y_{t}^{\gamma\left( d,t \right)}$ removes the bias from unmeasured confounding given $\gamma(d,t)$. Let $d=2$:

$$E\left[ Y_{t}^{\gamma\left( 2,t \right)}|D_{t}=2,\boldsymbol{X}_{t-1} \right]=E\left[ \left. \frac{\sum_{j=0}^{2} \left[ \Pr\left( D_{t}=j \right|\boldsymbol{X}_{t-1})Pr(Y_{t}\left( d=2 \right)=1|D_{t}=j,\boldsymbol{X}_{t-1}) \right]}{\Pr\left( Y_{t}\left( d=2 \right)=1 | D_{t}=2, \boldsymbol{X}_{t-1} \right)}Y_{t} \right|D_{t}=2, \boldsymbol{X}_{t-1} \right]= E\left[ \left. \frac{Y_{t}}{\Pr\left( Y_{t}\left( d=2 \right)=1 | D_{t}=2, \boldsymbol{X}_{t-1} \right)}Pr(Y_{t}\left( d=2 \right)=1|\boldsymbol{X}_{t-1}) \right|D_{t}=2, \boldsymbol{X}_{t-1} \right]=\frac{Pr(Y_{t}\left( d=2 \right)=1|\boldsymbol{X}_{t-1})}{Pr(Y_{t}\left( d=2 \right)=1|D_{t}=2, \boldsymbol{X}_{t-1})}E\left[ Y_{t} | D_{t}=2,\boldsymbol{X}_{t-1} \right]=\frac{Pr(Y_{t}\left( d=2 \right)=1|\boldsymbol{X}_{t-1})}{Pr(Y_{t}\left( d=2 \right)=1|D_{t}=2, \boldsymbol{X}_{t-1})}\Pr\left[ Y_{t}\left( d=2 \right)=1 | D_{t}=2,\boldsymbol{X}_{t-1} \right] by consistency =\Pr\left( Y_{t}\left( d=2 \right)=1 | \boldsymbol{X}_{t-1} \right).$$

The arguments for $d=0$ and $d=1$ are similar. Thus, the use of $Y_{t}^{\gamma(d,t)}$removes bias from unmeasured confounding given $\gamma(d,t)$.

In our implementation, we assumed that $\alpha_{1}\left( d, j \right)=\alpha_{2}\left( d, j \right)\equiv\alpha\left( d, j \right) \forall d, j$,

$$\frac{Pr(Y_{t}\left( d \right)=1|D_{t}=1,\boldsymbol{X}_{t-1})}{Pr(Y_{t}\left( d \right)=1|D_{t}=0, \boldsymbol{X}_{t-1})}=RR_{c},$$

$$\frac{\Pr\left( Y_{t}\left( d \right)=1 | D_{t}=2,\boldsymbol{X}_{t-1} \right)}{\Pr\left( Y_{t}\left( d \right)=1 | D_{t}=0, \boldsymbol{X}_{t-1} \right)}=RR_{c}^{2},$$

$$\frac{Pr(Y_{t}\left( d \right)=1|D_{t}=0,\boldsymbol{X}_{t-1})}{Pr(Y_{t}\left( d \right)=1|D_{t}=1, \boldsymbol{X}_{t-1})}=\frac{1}{RR_{c}},$$

$$\frac{Pr(Y_{t}\left( d \right)=1|D_{t}=2,\boldsymbol{X}_{t-1})}{Pr(Y_{t}\left( d \right)=1|D_{t}=1, \boldsymbol{X}_{t-1})}=RR_{c},$$

$$\frac{\Pr\left( Y_{t}\left( d \right)=1 | D_{t}=0,\boldsymbol{X}_{t-1} \right)}{\Pr\left( Y_{t}\left( d \right)=1 | D_{t}=2, \boldsymbol{X}_{t-1} \right)}=\frac{1}{RR_{c}^{2}},$$

and

$$\frac{Pr(Y_{t}\left( d \right)=1|D_{t}=1,\boldsymbol{X}_{t-1})}{Pr(Y_{t}\left( d \right)=1|D_{t}=2, \boldsymbol{X}_{t-1})}=\frac{1}{RR_{c}},$$

As with the $A$ analysis, we evaluated $RR_{c}\in\{0.5, 0.63, 0.71, 0.8, 1, 1.25, 1.4, 1.6, 2\}$.

**REFERENCES**

1.	Koblin, B.A., et al., *Correlates of HIV acquisition in a cohort of Black men who have sex with men in the United States: HIV prevention trials network (HPTN) 061.* PLOS One 2013. **8**(7): p. e70413.

2.	Wheeler, D.P., et al., *Pre‐exposure prophylaxis initiation and adherence among Black men who have sex with men (MSM) in three US cities: results from the HPTN 073 study.* Journal of the International AIDS Society, 2019. **22**(2): p. e25223.

3.	Hendrix, C.W., et al., *Dose frequency ranging pharmacokinetic study of tenofovir-emtricitabine after directly observed dosing in healthy volunteers to establish adherence benchmarks (HPTN 066).* AIDS Research and Human Retroviruses, 2016. **32**(1): p. 32-43.

4.	Wang, C., et al., *Propensity score-integrated composite likelihood approach for incorporating real-world evidence in single-arm clinical studies.* Journal of Biopharmaceutical Statistics, 2020. **30**(3): p. 495-507.

5.	Fang, Y., et al., *Key considerations in the design of real-world studies.* Contemporary Clinical Trials, 2020. **96**: p. 106091.

6.	McCulloch, C.E., *observational studies, time-dependent confounding, and marginal structural models.* Arthritis & Rheumatology, 2015. **67**(3): p. 609-611.

7.	Degtiar, I. and S. Rose, *A review of generalizability and transportability.* Annual Review of Statistics and Its Application, 2023. **10**: p. 501-524.

8.	Thorlund, K., et al., *Synthetic and external controls in clinical trials–a primer for researchers.* Clinical Epidemiology, 2020: p. 457-467.

9.	Imbens, G.W., *The role of the propensity score in estimating dose-response functions.* Biometrika, 2000. **87**(3): p. 706-710.

10.	Westreich, D., et al., *Imputation approaches for potential outcomes in causal inference.* International Journal of Epidemiology, 2015. **44**(5): p. 1731-1737.

11.	Daniel, R.M., et al., *Methods for dealing with time‐dependent confounding.* Statistics in Medicine, 2013. **32**(9): p. 1584-1618.

12.	Hernán, M.A. and J.M. Robins, *Estimating causal effects from epidemiological data.* Journal of Epidemiology & Community Health, 2006. **60**(7): p. 578-586.

13.	Greifer, N. and E.A. Stuart, *Choosing the estimand when matching or weighting in observational studies*. 2021: arXiv preprint arXiv:2106.10577.

14.	Hernán, M.A., B.A. Brumback, and J.M. Robins, *Estimating the causal effect of zidovudine on CD4 count with a marginal structural model for repeated measures.* Statistics in Medicine, 2002. **21**(12): p. 1689-1709.

15.	Naimi, A.I., S.R. Cole, and E.H. Kennedy, *An introduction to g methods.* International Journal of Epidemiology, 2017. **46**(2): p. 756-762.

16.	Hernán, M.Á., B. Brumback, and J.M. Robins, *Marginal structural models to estimate the causal effect of zidovudine on the survival of HIV-positive men.* Epidemiology, 2000: p. 561-570.

17.	Platt, R.W., J.A.C. Delaney, and S. Suissa, *The positivity assumption and marginal structural models: the example of warfarin use and risk of bleeding.* European Journal of Epidemiology, 2012. **27**: p. 77-83.

18.	Xiao, Y., M. Abrahamowicz, and E.E. Moodie, *Accuracy of conventional and marginal structural Cox model estimators: a simulation study.* The International Journal of Biostatistics, 2010. **6**(2).

19.	Yang, S., et al., *Application of marginal structural models in pharmacoepidemiologic studies: a systematic review.* Pharmacoepidemiology and Drug Safety, 2014. **23**(6): p. 560-571.

20.	Hernán, M.A., *Beyond exchangeability: the other conditions for causal inference in medical research.* Statistical Methods in Medical Research 2012. **21**(1): p. 3-5.

21.	Bacak, V. and E.H. Kennedy, *Marginal structural models: An application to incarceration and marriage during young adulthood.* Journal of Marriage and Family, 2015. **77**(1): p. 112-125.

22.	Richardson, T.S. and J.M. Robins, *Single world intervention graphs (SWIGs): A unification of the counterfactual and graphical approaches to causality.* Center for the Statistics and the Social Sciences, University of Washington Series. Working Paper, 2013. **128**(30): p. 2013.

23.	Ertefaie, A. and D.A. Stephens, *Comparing approaches to causal inference for longitudinal data: inverse probability weighting versus propensity scores.* The International Journal of Biostatistics, 2010. **6**(2).

24.	Li, F. and F. Li, *Propensity score weighting for causal inference with multiple treatments.* The Annals of Applied Statistics, 2019. **13**(4): p. 2389-2415.

25.	Hernán, M.A., *A definition of causal effect for epidemiological research.* Journal of Epidemiology & Community Health, 2004. **58**(4): p. 265-271.

26.	Robins, J.M., M.A. Hernan, and B. Brumback, *Marginal structural models and causal inference in epidemiology.* Epidemiology, 2000: p. 550-560.

27.	Cerdá, M., et al., *The relationship between neighborhood poverty and alcohol use: estimation by marginal structural models.* Epidemiology, 2010. **21**(4): p. 482.

28.	Shinozaki, T. and E. Suzuki, *Understanding marginal structural models for time-varying exposures: pitfalls and tips.* Journal of Epidemiology, 2020. **30**(9): p. 377-389.

29.	Howe, C.J., et al., *Estimating the effects of multiple time-varying exposures using joint marginal structural models: alcohol consumption, injection drug use, and HIV acquisition.* Epidemiology, 2012. **23**(4): p. 574.

30.	Stekhoven, D.J. and P. Bühlmann, *MissForest—non-parametric missing value imputation for mixed-type data.* Bioinformatics, 2012. **28**(1): p. 112-118.

31.	Mayer, M., *missRanger: Fast Imputation of Missing Values*. 2023. p. R package.

32.	Leyrat, C., et al., *Propensity score analysis with partially observed covariates: how should multiple imputation be used?* Statistical Methods in Medical Research, 2019. **28**(1): p. 3-19.

33.	Mattei, A., *Estimating and using propensity score in presence of missing background data: an application to assess the impact of childbearing on wellbeing.* Statistical Methods and Applications, 2009. **18**: p. 257-273.

34.	Schomaker, M. and C. Heumann, *Bootstrap inference when using multiple imputation.* Statistics in Medicine, 2018. **37**(14): p. 2252-2266.

35.	Austin, P.C. and E.A. Stuart, *Moving towards best practice when using inverse probability of treatment weighting (IPTW) using the propensity score to estimate causal treatment effects in observational studies.* Statistics in Medicine, 2015. **34**(28): p. 3661-3679.

36.	Austin, P.C., *An introduction to propensity score methods for reducing the effects of confounding in observational studies.* Multivariate Behavioral Research, 2011. **46**(3): p. 399-424.

37.	Fong, C. and K. Imai, *Covariate balancing propensity score for general treatment regimes*, in *Princeton Manuscript*. 2014.

38.	McCaffrey, D.F., et al., *A tutorial on propensity score estimation for multiple treatments using generalized boosted models.* Statistics in Medicine, 2013. **32**(19): p. 3388-3414.

39.	Greifer, N., *cobalt: Covariate Balance Tables and Plots*. 2023. p. R package version.

40.	Xiao, Y., E.E. Moodie, and M. Abrahamowicz, *Comparison of approaches to weight truncation for marginal structural Cox models.* Epidemiologic Methods, 2013. **2**(1): p. 1-20.

41.	Naimi, A.I., et al., *A comparison of methods to estimate the hazard ratio under conditions of time-varying confounding and nonpositivity.* Epidemiology (2011. **22**(5): p. 718.

42.	Westreich, D. and S.R. Cole, *Invited commentary: positivity in practice.* American Journal of Epidemiology, 2010. **171**(6): p. 674-677.

43.	Barber, J.S., S.A. Murphy, and N. Verbitsky, *Adjusting for time-varying confounding in survival analysis.* Sociological Methodology, 2004. **34**(1): p. 163-192.

44.	Robins, J.M., *Association, causation, and marginal structural models.* Synthese, 1999. **121**(1/2): p. 151-179.

45.	Brumback, B.A., et al., *Sensitivity analyses for unmeasured confounding assuming a marginal structural model for repeated measures.* Statistics in Medicine, 2004. **23**(5): p. 749-767.

46.	Chiba, Y., *Sensitivity analysis of unmeasured confounding for the causal risk ratio by applying marginal structural models.* Communications in Statistics - Theory and Methods, 2009. **39**(1): p. 65-76.

47.	Klungsøyr, O., et al., *Sensitivity analysis for unmeasured confounding in a marginal structural Cox proportional hazards model.* Lifetime Data Analysis, 2009. **15**: p. 278-294.

48.	Hernán, M.A. and T.J. VanderWeele, *Compound treatments and transportability of causal inference.* Epidemiology, 2011. **22**(3): p. 368.

49.	VanderWeele, T.J. and M.J. Knol, *A tutorial on interaction.* Epidemiologic Methods, 2014. **3**(1): p. 33-72.

50.	Słoczyński, T., *Interpreting OLS estimands when treatment effects are heterogeneous: Smaller groups get larger weights.* Review of Economics and Statistics, 2022. **104**(3): p. 501-509.

51.	Goedel, W.C., et al., *A pilot study of a patient navigation intervention to improve HIV pre-exposure prophylaxis persistence among black/African American men who have sex with men.* JAIDS Journal of Acquired Immune Deficiency Syndromes, 2022: p. 10.1097.

52.	Cole, S.R., et al., *Effect of highly active antiretroviral therapy on time to acquired immunodeficiency syndrome or death using marginal structural models.* American Journal of Epidemiology, 2003. **158**(7): p. 687-694.

53.	Hernán, M.A., B. Brumback, and J.M. Robins, *Marginal structural models to estimate the joint causal effect of nonrandomized treatments.* Journal of the American Statistical Association, 2001. **96**(454): p. 440-448.

54.	Burcu, M., et al., *Real‐world evidence to support regulatory decision‐making for medicines: considerations for external control arms.* Pharmacoepidemiology and Drug Safety, 2020. **29**(10): p. 1228-1235.

55.	Westreich, D., et al., *Transportability of trial results using inverse odds of sampling weights.* American Journal of Epidemiology, 2017. **186**(8): p. 1010-1014.

56.	Molina, J.-M., et al., *On-demand preexposure prophylaxis in men at high risk for HIV-1 infection.* New England Journal of Medicine, 2015. **373**(23): p. 2237-2246.

57.	Dimitrov, D.T., B.R. Mâsse, and D. Donnell, *PrEP adherence patterns strongly impact individual HIV risk and observed efficacy in randomized clinical trials.* JAIDS Journal of Acquired Immune Deficiency Syndromes, 2016. **72**(4): p. 444.

58.	Donnell, D., et al., *HIV protective efficacy and correlates of tenofovir blood concentrations in a clinical trial of PrEP for HIV prevention.* JAIDS: Journal of Acquired Immune Deficiency Syndromes, 2014. **66**(3): p. 340.

59.	Landovitz, R.J., et al., *Cabotegravir for HIV prevention in cisgender men and transgender women.* New England Journal of Medicine, 2021. **385**(7): p. 595-608.

60.	Von Hippel, P.T., *4. Regression with missing Ys: an improved strategy for analyzing multiply imputed data.* Sociological Methodology, 2007. **37**(1): p. 83-117.

61.	Sullivan, T.R., et al., *Bias and precision of the “multiple imputation, then deletion” method for dealing with missing outcome data.* American Journal of Epidemiology, 2015. **182**(6): p. 528-534.

62.	Jakobsen, J.C., et al., *When and how should multiple imputation be used for handling missing data in randomised clinical trials–a practical guide with flowcharts.* BMC Medical Research Methodology, 2017. **17**(1): p. 1-10.

63.	Radloff, L.S., *The CES-D scale: A self-report depression scale for research in the general population.* Applied Psychological Measurement, 1977. **1**(3): p. 385-401.

64.	Bogart, L.M. and S. Thorburn, *Are HIV/AIDS conspiracy beliefs a barrier to HIV prevention among African Americans?* JAIDS Journal of Acquired Immune Deficiency Syndromes, 2005. **38**(2): p. 213-218.

65.	Brooks, R.A., et al., *HIV/AIDS conspiracy beliefs and intention to adopt preexposure prophylaxis among black men who have sex with men in Los Angeles.* International Journal of STD & AIDS, 2018. **29**(4): p. 375-381.

66.	Herek, G. and E.K. Glunt, *Identity and community among gay and bisexual men in the AIDS era*, in *AIDS, Identity, and Community*, G. Herek and B. Greene, Editors. 1995, SAGE. p. 55.

67.	Berkman, L.F. and S.L. Syme, *Social networks, host resistance, and mortality: a nine-year follow-up study of Alameda County residents.* American Journal of Epidemiology, 1979. **109**(2): p. 186-204.

68.	Sayles, J.N., et al., *Development and psychometric assessment of a multidimensional measure of internalized HIV stigma in a sample of HIV-positive adults.* AIDS and Behavior, 2008. **12**: p. 748-758.

69.	Saunders, J.B., et al., *Development of the alcohol use disorders identification test (AUDIT): WHO collaborative project on early detection of persons with harmful alcohol consumption‐II.* Addiction, 1993. **88**(6): p. 791-804.

70.	Sato, T. and Y. Matsuyama, *Marginal structural models as a tool for standardization.* Epidemiology, 2003: p. 680-686.
